# Comparing the risks of new-onset gastric cancer or gastric diseases in type 2 diabetes mellitus patients exposed to SGLT2I, DPP4I or GLP1A: a population-based cohort study

**DOI:** 10.1101/2023.07.02.23292063

**Authors:** Oscar Hou In Chou, Vinod Kumar Chauhan, Cheuk To Chung, Lei Lu, Teddy Tai Loy Lee, Zita Man Wai Ng, Karin Kai Wang, Sharen Lee, Haipeng Liu, Wing Tak Wong, Ronald Ting Kai Pang, Apichat Kaewdech, Bernard Man Yung Cheung, Gary Tse, Jiandong Zhou

## Abstract

**Objective:** To compare the risks of gastric cancer and other gastric diseases upon exposed to sodium-glucose cotransporter 2 inhibitors (SGLT2I), dipeptidyl peptidase-4 inhibitors (DPP4I) and glucagon-like peptide-1 receptor agonists (GLP1A).

**Design:** This was a retrospective population-based cohort study of prospectively recorded data on type-2 diabetes mellitus (T2DM) patients prescribed either SGLT2I or DPP4I between January 1^st^ 2015 and December 31^st^ 2020 from Hong Kong. The primary outcome was new-onset gastric cancer, peptic ulcer (PU), acute gastritis, non-acute gastritis, and gastroesophageal reflux disease (GERD). Propensity score matching (1:1 ratio) using the nearest neighbour search was performed and multivariable Cox regression was carried out. A three-arm analysis including the GLP1A cohort was subsequently conducted.

**Results:** A total of 62858 T2DM patients (median age: 62.2 years old [SD: 12.8]; 55.93% males; SGLT2I: n=23442; DPP4I: n=39416) were included. After matching, the incidence of gastric cancer was significantly lower in SGLT2I users (Incidence rate, IR: 0.32; 95% confidence interval, CI: 0.23-0.43) than DPP4I users (IR: 1.22; CI: 1.03-1.42). SGLT2I use was associated with lower risks of gastric cancer (HR: 0.30; 95% CI: 0.19-0.48) after adjusting for significant covariates compared to DPP4I use. SGLT2 use was also associated with lower risks of PU, acute gastritis, non-acute gastritis, and GERD (all p<0.05). The three-arm analysis demonstrated higher risks of gastric cancer and GERD in GLP1A than in SGLT2I.

**Conclusions:** SGLT2I was associated with lower risks of new-onset gastric cancer, PU, acute gastritis, non-acute gastritis, and GERD after matching and adjustments compared to DPP4I.

**Graphical abstract:** 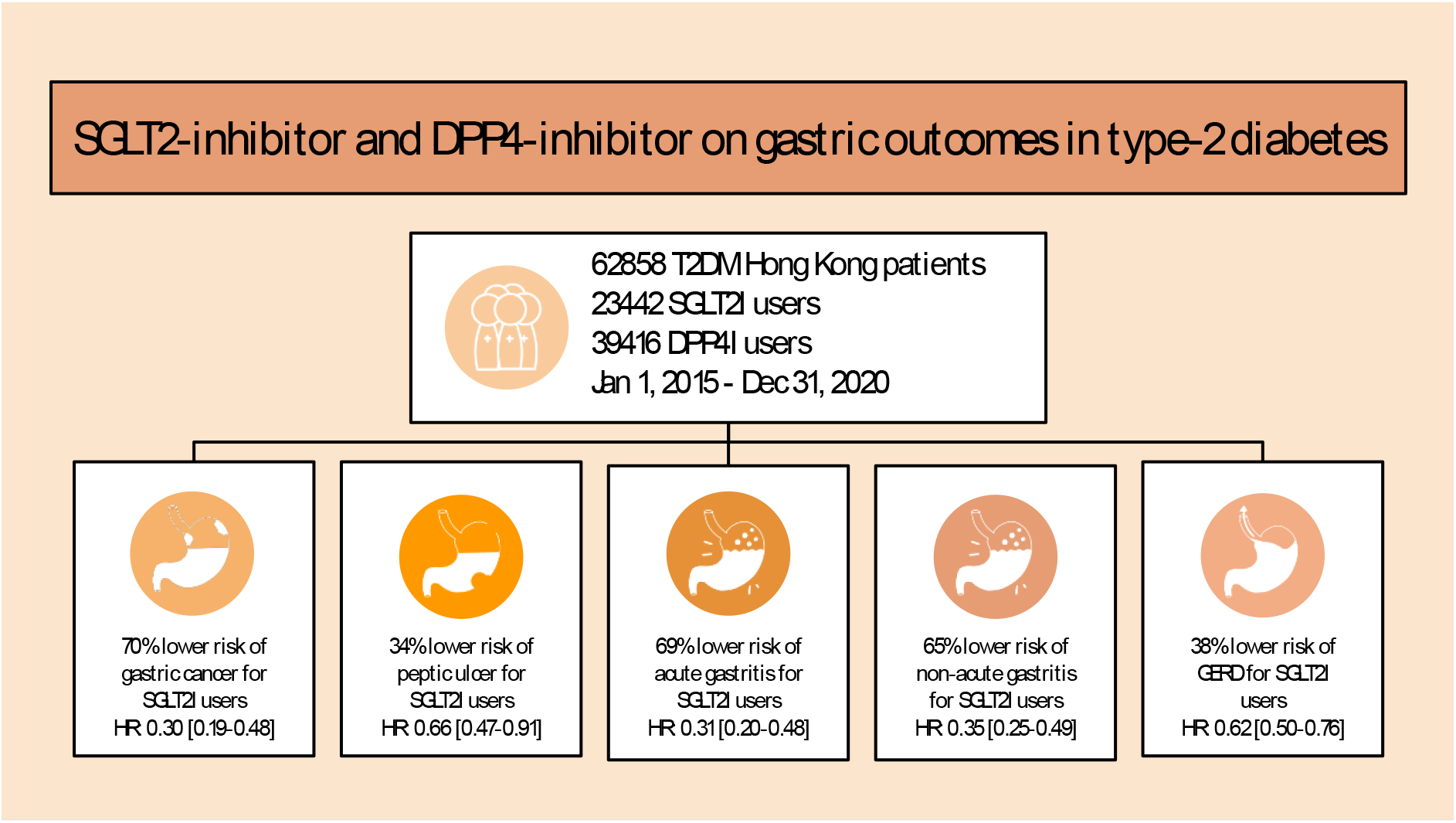

**Key messages:** 

**What is already known on this topic:** T2DM was associated with higher risks of gastric cancer. Anti-diabetic drugs such as SGLT2I and DPP4I are commonly used second-line drugs to help manage diabetes mellitus. SGLT2I was previously suggested to lower the risks of cancer compared to DPP4I.

**What this study adds:** In this population-based cohort study, SGLT2I was associated with a 70% lower risk of gastric cancer, 34% low risks of peptic ulcer, 69% lower risks of acute gastritis, 65% low risks of non-acute gastritis, and 38% of gastroesophageal reflux disease (GERD) than DPP4I users. The three arm sensitivity analysis involving DPP4I and GLP1A using stabilized inverse probability treatment weighting demonstrated higher risks of gastric cancer and GERD in GLP1A users compared to SGLT2I users.

**How this study might affect research, practice or policy:** The findings of this study may influence the choice of novel second-line anti-diabetic therapy in T2DM patients in terms of the gastric safety profile especially gastric cancer. This study may inspire further mechanistic studies and studies on the long-term cancer benefits of SGLT2I.

## Introduction

The rising incidence of type 2 diabetes mellitus (T2DM) is a major global concern, imposing a significant burden on healthcare systems worldwide. T2DM is associated with several complications, in particular, T2DM has been linked to increased upper gastrointestinal (GI) tract complications. Gastric cancer is one of the most common causes of cancer-related mortality, estimating a 5-year overall survival rate of less than 30% [1]. A systematic review and meta-analysis of cohort studies has revealed a 14% higher risk of gastric cancer in individuals with T2DM [2]. Gastric cancer is significantly more prevalent in Asia, with about half of the total gastric cancer cases found in the region, especially in China [3, 4].

Besides of gastric cancer, there is growing evidence regarding the role of diabetes in various types of gastric diseases. Patients with diabetes were found to be at higher risk for developing peptic ulcers [5]. In addition to this, a case-control study revealed that diabetic patients had a higher propensity to atrophic corpus gastritis, a significant risk factor for distal gastric cancer [6]. Therefore, the evidences raised the question whether anti-diabetic agents had protective effects against gastric diseases, especially gastric cancer. Previous studies demonstrated the effects of metformin on healing gastric ulcers and reducing risks of gastric cancer [7]. For instance, it was demonstrated that metformin was associated with decreased risk in gastric cancer among *Helicobacter pylori (H. pylori)*-eradicated diabetic patients [8].

However, the current literature surrounding novel anti-diabetic agents such as sodium-glucose cotransporter 2 inhibitors (SGLT2I) and dipeptidyl peptidase 4 inhibitors (DPP4I) on various gastric disease remain controversial. Recent studies suggested that SGLT2I may offer potential gastrointestinal protective effects. An experimental study has shown that empagliflozin can increase the production of gastric mucin, exerting protective effects against gastritis in animal models [9]. Furthermore, a meta-analysis of randomized controlled trials demonstrated an association between canagliflozin and a decreased risk of GI cancers, but no significant association was found with other SGLT2Is [10]. However, there is a lack of clinical studies focusing on the effects of DPP4Is on the gastric outcomes of patients with T2DM. As of now, there is limited clinical evidence surrounding the association of SGLT2I and DPP4I on different types of gastric diseases. Hence, the present study aims to explore the role of SGLT2I and DPP4I on new-onset gastric cancer and gastric diseases in a cohort of T2DM patients from Hong Kong.

## Methods

### Study population

This was a retrospective population-based study of prospectively collected patient records using the Clinical Data Analysis and Reporting System (CDARS). This system hosts data on disease diagnosis, laboratory results, past comorbidities, clinical characteristics and medication prescriptions. The system has been used by our Hong Kong research team to perform comparative studies [11, 12, 13]. This study was approved by the Institutional Review Board of the University of Hong Kong/Hospital Authority Hong Kong West Cluster and The Joint Chinese University of Hong Kong-New Territories East Cluster, and complied with the Declaration of Helsinki. In this study, patients who were diagnosed with T2DM and were administered either only SGLT2I or only DPP4I in centres under the Hong Kong Hospital Authority, between January 1st, 2015 and December 31st, 2020 were included. These cohorts have been used by previously by us to compare the effects between SGLT2I and DPP4I on cardiovascular [14, 15], neurological [16] and cancer outcomes [17]. The glucagon-like peptide-1 receptor agonist (GLP1A) cohort was included for additional comparisons.

### Predictors and variables

Patients’ demographics include gender and age of initial drug use (baseline), clinical and biochemical data were extracted for the present study. Prior comorbidities were extracted in accordance with the *International Classification of Diseases Ninth Edition* (ICD-9) codes (**Supplementary Table 1**). Patient with prior identified *H. pylori* infection were defined by either ICD-9: 041.86, Microbiology results (MIS data), or prior history of *H. pylori* eradication therapy with concomitant use of proton pump inhibitor (PPI) and antibiotics according to previous study [18]. The diabetes duration was calculated by examining the earliest date amongst the first date of (1) diagnosis using of ICD-9; (2) Haemoglobin A1c (HbA1c) ≥6.5%; (3) Fasting glucose ≥7.0 mmol/l or Random glucose ≥11.1 mmol/l 4) Using anti-diabetic medications. The Charlson’s standard comorbidity index was calculated. Patients with hepatitis C virus (HCV) infection were defined by both the ICD-9 code and positive HCV-RNA positive status. Patients with hepatitis B virus (HBV) infection were defined by both the ICD-9 code and positive hepatitis B surface antigen (HBsAg) positive status.

The GI medications, cardiovascular medications, anti-diabetic agents, and the antibiotics were extracted from the database. The duration and frequency of SGLT2I and DPP4I usage were extracted. Moreover, the baseline laboratory examinations, including the complete blood count, lipid and glucose profiles, as well as liver and renal test results were extracted. The estimated glomerular filtration rate (eGFR) was calculated using the abbreviated modification of diet in renal disease (MDRD) equation [19]. Different metrics of variability in lipid and glucose tests were also calculated to reflect their fluctuations **(Supplementary Table 2)**.

### Adverse outcomes

The primary outcome of this study was the development of gastric cancer, peptic ulcer, acute gastritis, non-acute gastritis, and gastroesophageal reflux disease (GERD) upon the index date of the drug use **(Supplementary Table 1)**. The secondary outcome was all-cause mortality as a sensitivity analysis. Mortality data were obtained from the Hong Kong Death Registry, a population-based official government registry with the registered death records of all Hong Kong citizens linked to CDARS. Mortality was recorded using the *International Classification of Diseases Tenth Edition* (ICD-10) coding. The endpoint date of interest for eligible patients was the event presentation date. The endpoint for those without primary outcome presentation was the mortality date or the endpoint of the study (31^st^ December 2020).

### Statistical analysis

Descriptive statistics are used to summarize baseline clinical and biochemical characteristics of patients with SGLT2I and DPP4I use. For baseline clinical characteristics, continuous variables were presented as mean (95% confidence interval [95% CI]/standard deviation [SD]) and the categorical variables were presented as total numbers (percentage). Propensity score matching with 1:1 ratio for SGLT2I use versus DPP4I use based on demographics, Charlson comorbidity index, non-SGLT2I/DPP4I medications, prior comorbidities, biomarkers and duration from T2DM diagnosis initial drug exposure were performed using the nearest neighbour search strategy with a calliper of 0.1. Propensity score matching was performed using Stata software (Version 16.0).

Baseline characteristics between patients with SGLT2I and DPP4I use before and after matching were compared with absolute standardized mean difference (SMD), with SMD<0.10 regarded as well-balanced between the two groups. The cumulative incidence curves for the primary outcomes and secondary outcomes were constructed and compared for the risk using log-rank tests. Proportional Cox regression models were used to identify significant risk predictors of adverse study outcomes. Subgroup analysis was conducted to confirm the association amongst patients with different clinical important predictors.

Cause-specific and sub-distribution hazard models were conducted to consider possible competing risks. Multiple propensity adjustment approaches were used, including propensity score stratification [20], propensity score matching with inverse probability of treatment weighting [21] and propensity score matching with stable inverse probability weighting [22]. The three arm sensitivity results involving GLP1A using stabilized IPTW were conducted to confirm the association and choice of second-line anti-diabetic medications. Furthermore, patients with chronic kidney disease (CKD) stage 4/5 (eGFR <30), peritoneal dialysis or haemodialysis who may be contraindicated with SGLT2I were excluded in the sensitivity analysis. Sensitivity analysis results with consideration of one-year lag time effects was also conducted. The as-treat approach was conducted, which patient were censored at treatment discontinuation or switching of the comparison medications. The hazard ratio (HR), 95% CI and P-value were reported. Statistical significance was defined as P-value <0.05. All statistical analyses were performed with RStudio (Version: 1.1.456) and Python (Version: 3.6).

## Results

### Basic characteristics

In this territory-wide cohort study of 76147 patients with T2DM treated with SGLT2I/DPP4I between 1^st^ January 2015 and 31^st^ December 2020 in Hong Kong, patients were followed up until 31^st^ December 2020 or until their deaths (**Figure 1**). The following patient groups were excluded: those who (1) died within 30 days after initial drug exposure (N=167); (2) without complete demographics (N=19); (3) under 18 years old (N=108); (4) with prior peptic ulcer, gastritis, GERD, gastric cancer (N=108); (5) exposed to both DPP4I and SGLT2I prescription at any time point (N=12858) and (6) new onset gastric cancer development less than 1 year after drug exposure (N=29).

**Figure 1.**
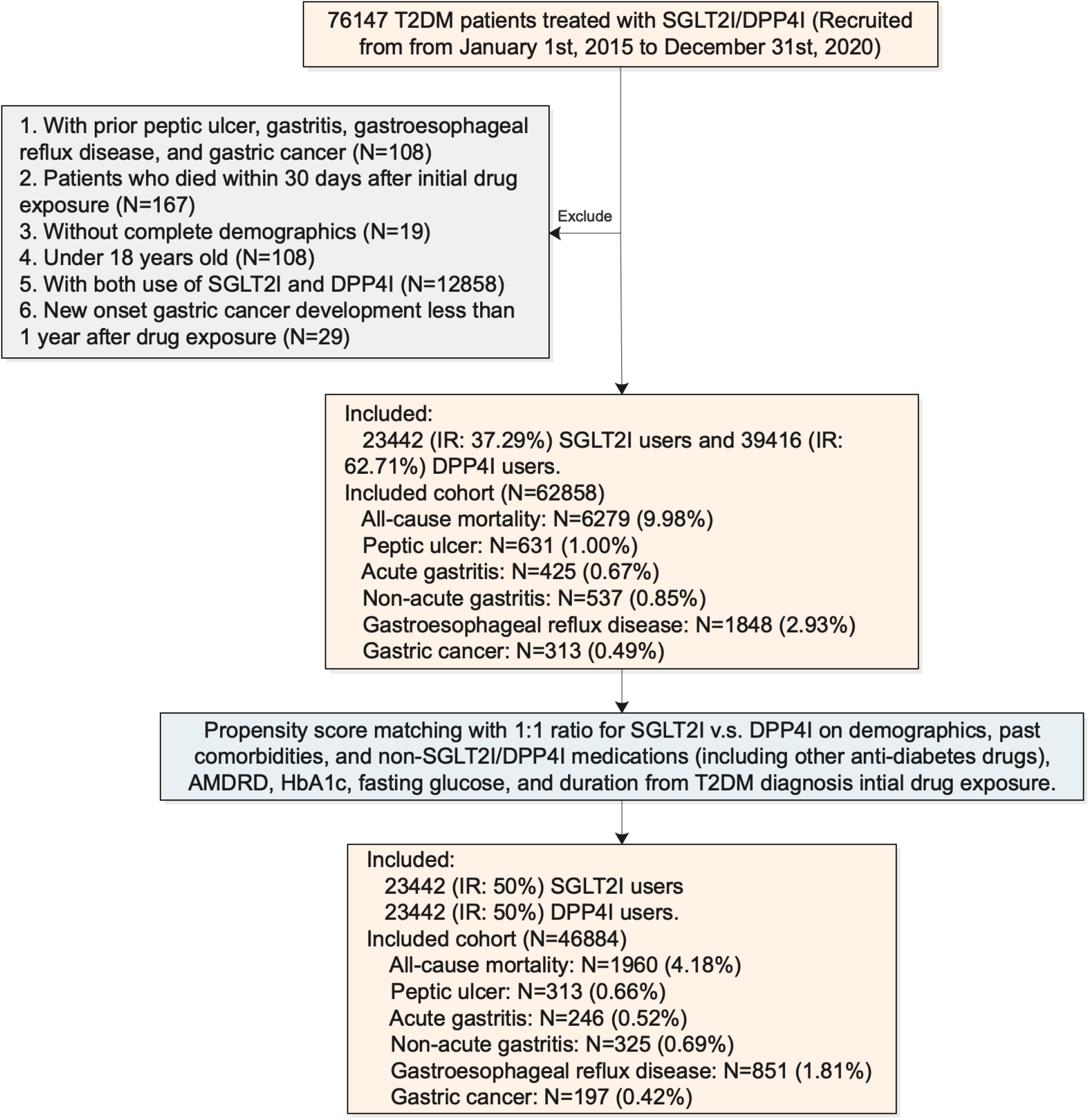
Procedures of data processing for the study cohort SGLT2I: Sodium-glucose cotransporter-2 inhibitors; DPP4I: Dipeptidyl peptidase-4 inhibitors. MDRD: modification of diet in renal disease.

After exclusion, this study included a total of 62858 patients with T2DM (mean age: 62.2 years old [SD: 12.8]; 55.93% males), of whom 23442 patients (35.33%) used SGLT2Is, and 39416 patients (64.67%) used DPP4Is (**Table 1**). Before matching, the SGLT2 users were younger, with less comorbidities, and more patients using anti-diabetic drugs thiazolidinedione and GLP1A, have a higher eGFR compared to DPP4I users.

**Table 1.** Baseline and clinical characteristics of patients with SGLT2I v.s. DPP4I use before and after propensity score matching (1:1).

| Characteristics | Before matching |  |  | SMD<sup>#</sup> |
| --- | --- | --- | --- | --- |
|  | All (N=62858)<br>Mean(SD);N or<br>Count(%) | SGLT2I users<br>(N=23442)<br>Mean(SD);N or<br>Count(%) | DPP4I users<br>(N=39416)<br>Mean(SD);N or<br>Count(%) |  |
| Demographics |  |  |  |  |
| Male gender | 35160(55.93%) | 13979(59.63%) | 21181(53.73%) | 0.12 |
| Female gender | 27698(44.06%) | 9463(40.36%) | 18235(46.26%) | 0.12 |
| Baseline age, years | 62.2(12.8);n=62858 | 57.7(11.3);n=23442 | 64.9(12.9);n=39416 | 0.59* |
| Past comorbidities |  |  |  |  |
| Charlson comorbidity index | 2.0(1.5);n=62858 | 1.6(1.3);n=23442 | 2.3(1.6);n=39416 | 0.52* |
| Duration from earliest diabetes mellitus diagnosis to index date, day | 509.9(1214.8);n=62858 | 519.6(1332.2);n=23442 | 504.1(1139.2);n=39416 | 0.01 |
| Hypertension | 14514(23.09%) | 5255(22.41%) | 9259(23.49%) | 0.03 |
| Heart failure | 2080(3.30%) | 634(2.70%) | 1446(3.66%) | 0.05 |
| Ischemic heart disease | 6381(10.15%) | 2904(12.38%) | 3477(8.82%) | 0.12 |
| Atrial fibrillation | 1604(2.55%) | 526(2.24%) | 1078(2.73%) | 0.03 |
| Acute myocardial infarction | 1775(2.82%) | 798(3.40%) | 977(2.47%) | 0.05 |
| Peripheral vascular disease | 588(0.93%) | 193(0.82%) | 395(1.00%) | 0.02 |
| Renal diseases | 1140(1.81%) | 151(0.64%) | 989(2.50%) | 0.15 |
| Stroke/transient ischemic attack | 1989(3.16%) | 615(2.62%) | 1374(3.48%) | 0.05 |
| Gastrointestinal bleeding | 1434(2.28%) | 414(1.76%) | 1020(2.58%) | 0.06 |
| Identified H. pylori infection | 3597(5.72%) | 1231(5.25%) | 2366(6.00%) | 0.03 |
| Autoimmune disease | 653(1.03%) | 246(1.04%) | 407(1.03%) | <0.01 |
| Cancer | 1604(2.55%) | 448(1.91%) | 1156(2.93%) | 0.07 |
| Chronic liver disease and cirrhosis | 1312(2.08%) | 633(2.70%) | 679(1.72%) | 0.07 |
| HBV infection | 2640(4.19%) | 1232(5.25%) | 1408(3.57%) | 0.08 |
| HCV infection | 191(0.30%) | 70(0.29%) | 121(0.30%) | <0.01 |
| Other liver diseases | 711(1.13%) | 222(0.94%) | 489(1.24%) | 0.03 |
| Diabetic retinopathy | 4316(6.86%) | 1626(6.93%) | 2690(6.82%) | <0.01 |
| Medication prescriptions |  |  |  |  |
| SGLT2I frequency | 7.2(9.6);n=2344<br>2 | 7.2(9.6);n=2344<br>2 | - | - |
| DPP4I frequency | 5.5(7.7);n=3941<br>6 | - | 5.5(7.7);n=3941<br>6 | - |
| SGLT2I duration, days | 505.2(624.7);n=23442 | 505.2(624.7);n=23442 | - | - |
| DPP4I duration, days | 520.3(301.1);n=39416 | - | 520.3(301.1);n=39416 | - |
| Metformin | 56412(89.74%) | 21884(93.35%) | 34528(87.59%) | 0.2 |
| Sulphonylurea | 48382(76.97%) | 16902(72.10%) | 31480(79.86%) | 0.18 |
| Insulin | 31798(50.58%) | 11950(50.97%) | 19848(50.35%) | 0.01 |
| Acarbose | 1676(2.66%) | 890(3.79%) | 786(1.99%) | 0.11 |
| Thiozolidinedone | 12537(19.94%) | 6377(27.20%) | 6160(15.62%) | 0.29* |
| Glucagon-like peptide-1 receptor agonists | 2017(3.20%) | 1561(6.65%) | 456(1.15%) | 0.29* |
| H2 antagonists | 29352(46.69%) | 10764(45.91%) | 18588(47.15%) | 0.02 |
| Proton pump inhibitors | 19309(30.71%) | 7007(29.89%) | 12302(31.21%) | 0.03 |
| ACEI/ARB | 20586(32.75%) | 13846(59.06%) | 6740(17.09%) | 0.96* |
| Antibiotics | 41546(66.09%) | 14883(63.48%) | 26663(67.64%) | 0.09 |
| Other antihypertensive drugs | 2709(4.30%) | 2117(9.03%) | 592(1.50%) | 0.34* |
| Anti-hepatitis | 946(1.50%) | 466(1.98%) | 480(1.21%) | 0.06 |
| Statins and fibrates | 32843(52.24%) | 17296(73.78%) | 15547(39.44%) | 0.74* |
| HCV treatment | 746(1.18%) | 363(1.54%) | 383(0.97%) | 0.05 |
| HBV treatment | 898(1.42%) | 440(1.87%) | 458(1.16%) | 0.06 |
| Antiplatelets | 11502(18.29%) | 7429(31.69%) | 4073(10.33%) | 0.54* |
| Lipid-lowering drugs | 22612(35.97%) | 15358(65.51%) | 7254(18.40%) | 1.09* |
| Non-steroidal anti-inflammatory drugs | 11041(17.56%) | 7122(30.38%) | 3919(9.94%) | 0.53* |
| Diuretics | 11823(18.80%) | 7196(30.69%) | 4627(11.73%) | 0.48* |
| Beta-blockers | 9273(14.75%) | 5973(25.47%) | 3300(8.37%) | 0.47* |
| Calcium channel blockers | 16249(25.85%) | 10218(43.58%) | 6031(15.30%) | 0.65* |
| Subclinical biomarkers |  |  |  |  |
| Abbreviated MDRD, mL/min/1.73m<sup>2</sup> | 80.9(28.5);n=51653 | 90.0(24.2);n=19466 | 75.4(29.5);n=32187 | 0.54* |
| Neutrophil-to-lymphocyte ratio | 3.5(4.6);n=26410 | 3.0(3.6);n=10521 | 3.9(5.2);n=15889 | 0.2 |
| Red cell count, x10<sup>12</sup>/L | 4.5(0.7);n=32370 | 4.7(0.6);n=13137 | 4.4(0.7);n=19233 | 0.51* |
| Liver and renal functions |  |  |  |  |
| Urea, mmol/L | 6.5(3.5);n=51510 | 5.7(2.1);n=19426 | 7.0(4.0);n=32084 | 0.4* |
| Creatinine, umol/L | 93.8(74.2);n=51653 | 78.9(29.2);n=19466 | 102.8(90.0);n=32187 | 0.36* |
| Bilirubin, umol/L | 11.3(7.0);n=40057 | 11.5(6.2);n=16403 | 11.1(7.5);n=23654 | 0.05 |
| Lipid/glucose profiles |  |  |  |  |
| Triglyceride, mmol/L | 1.7(1.6);n=48474 | 1.8(1.9);n=18581 | 1.7(1.4);n=29893 | 0.08 |
| Low-density lipoprotein, mmol/L | 2.4(0.8);n=47587 | 2.39(0.81);n=18228 | 2.39(0.8);n=29359 | 0.01 |
| High-density lipoprotein, mmol/L | 1.2(0.3);n=48395 | 1.16(0.31);n=18544 | 1.21(0.34);n=29851 | 0.14 |
| Total cholesterol, mmol/L | 4.3(1.0);n=48526 | 4.35(1.03);n=18609 | 4.34(0.99);n=29917 | 0.01 |
| Baseline-Fasting glucose, mmol/L | 9.0(3.4);n=62858 | 9.2(3.2);n=23442 | 8.9(3.4);n=39416 | 0.09 |
| Baseline-Hemoglobin<br>A1C, % | 8.1(1.4);n=6285<br>8 | 8.3(1.4);n=2344<br>2 | 8.0(1.4);n=3941<br>6 | 0.19 |

\* for $SMD \geq 0.1$ ; SGLT2I: sodium glucose cotransporter-2 inhibitor; DPP4I: dipeptidyl peptidase-4 inhibitor; SD: standard deviation; RMS: Rooted mean square; CV: Coefficient of variation; MDRD: modification of diet in renal disease; ACEI: angiotensin-converting enzyme inhibitors; ARB: angiotensin II receptor blockers; HBV: hepatitis B virus; HCV: hepatitis C virus; SMD: standardized mean difference

| After matching |  |  |  |
| --- | --- | --- | --- |
| All (N=46884)<br>Mean(SD);N or<br>Count(%) | SGLT2I users<br>(N=23442)<br>Mean(SD);N or<br>Count(%) | DPP4I users<br>(N=23442)<br>Mean(SD);N or<br>Count(%) | SMD<sup>#</sup> |
| 27992(59.70%) | 13979(59.63%) | 14013(59.77%) | <0.01 |
| 18892(40.29%) | 9463(40.36%) | 9429(40.22%) | <0.01 |
| 58.0(11.1);n=46884 | 57.7(11.3);n=23442 | 58.4(10.9);n=23442 | 0.06 |
| 1.6(1.2);n=46884 | 1.56(1.25);n=23442 | 1.58(1.23);n=23442 | 0.02 |
| 507.8(1254.5);n=46884 | 519.6(1332.2);n=23442 | 495.9(1171.4);n=23442 | 0.02 |
| 10149(21.64%) | 5255(22.41%) | 4894(20.87%) | 0.04 |
| 1242(2.64%) | 634(2.70%) | 608(2.59%) | 0.01 |
| 5575(11.89%) | 2904(12.38%) | 2671(11.39%) | 0.03 |
| 1045(2.22%) | 526(2.24%) | 519(2.21%) | <0.01 |
| 1565(3.33%) | 798(3.40%) | 767(3.27%) | 0.01 |
| 384(0.81%) | 193(0.82%) | 191(0.81%) | <0.01 |
| 301(0.64%) | 151(0.64%) | 150(0.63%) | <0.01 |
| 1221(2.60%) | 615(2.62%) | 606(2.58%) | <0.01 |
| 815(1.73%) | 414(1.76%) | 401(1.71%) | <0.01 |
| 2388(5.09%) | 1231(5.25%) | 1157(4.93%) | 0.01 |
| 480(1.02%) | 246(1.04%) | 234(0.99%) | 0.01 |
| 894(1.90%) | 448(1.91%) | 446(1.90%) | <0.01 |
| 1245(2.65%) | 633(2.70%) | 612(2.61%) | 0.01 |
| 2365(5.04%) | 1232(5.25%) | 1133(4.83%) | 0.02 |
| 140(0.29%) | 70(0.29%) | 70(0.29%) | <0.01 |
| 441(0.94%) | 222(0.94%) | 219(0.93%) | <0.01 |
| 3038(6.47%) | 1626(6.93%) | 1412(6.02%) | 0.04 |
| 7.2(9.6);n=23442 | 7.2(9.6);n=23442 | - | - |
| 6.7(8.2);n=23442 | - | 6.7(8.2);n=23442 | - |
| 505.2(624.7);n=23442 | 505.2(624.7);n=23442 | - | - |
| 592.9(361.4);n=23442 | - | 592.9(361.4);n=23442 | - |
| 43866(93.56%) | 21884(93.35%) | 21982(93.77%) | 0.02 |
| 34646(73.89%) | 16902(72.10%) | 17744(75.69%) | 0.08 |
| 24030(51.25%) | 11950(50.97%) | 12080(51.53%) | 0.01 |
| 1581(3.37%) | 840(3.58%) | 741(3.16%) | 0.02 |
| 12007(25.61%) | 6377(27.20%) | 5630(24.01%) | 0.07 |
| 1951(4.16%) | 1061(4.52%) | 890(3.79%) | 0.04 |
| 21747(46.38%) | 10764(45.91%) | 10983(46.85%) | 0.02 |
| 13720(29.26%) | 7007(29.89%) | 6713(28.63%) | 0.03 |
| 20270(43.23%) | 10146(43.28%) | 10124(43.18%) | <0.01 |
| 30127(64.25%) | 14883(63.48%) | 15244(65.02%) | 0.03 |
| 2688(5.73%) | 1517(6.47%) | 1171(4.99%) | 0.06 |
| 923(1.96%) | 466(1.98%) | 457(1.94%) | <0.01 |
| 32519(69.36%) | 16696(71.22%) | 15823(67.49%) | 0.08 |
| 723(1.54%) | 363(1.54%) | 360(1.53%) | <0.01 |
| 878(1.87%) | 440(1.87%) | 438(1.86%) | <0.01 |
| 32880(70.13%) | 16440(70.13%) | 16440(70.13%) | <0.01 |
| 11382(24.27%) | 5829(24.86%) | 5553(23.68%) | 0.03 |
| 20875(44.52%) | 10358(44.18%) | 10517(44.86%) | 0.01 |
| 10721(22.86%) | 5422(23.12%) | 5299(22.60%) | 0.01 |
| 11021(23.50%) | 5596(23.87%) | 5425(23.14%) | 0.02 |
| 9052(19.30%) | 4573(19.50%) | 4479(19.10%) | 0.01 |
| 88.4(24.8);n=37875 | 90.0(24.2);n=19466 | 86.8(25.3);n=18409 | 0.13* |
| 3.0(3.6);n=20694 | 3.0(3.6);n=10521 | 3.1(3.6);n=10173 | 0.03 |
| 4.7(0.6);n=25159 | 4.74(0.59);n=13137 | 4.68(0.61);n=12022 | 0.09 |
| 5.8(2.3);n=37790 | 5.7(2.1);n=19426 | 5.9(2.6);n=18364 | 0.09 |
| 81.5(41.1);n=37875 | 78.9(29.2);n=19466 | 84.1(50.6);n=18409 | 0.12 |
| 11.4(6.1);n=31503 | 11.5(6.2);n=16403 | 11.4(5.9);n=15100 | 0.02 |
| 1.8(1.8);n=36107 | 1.83(1.86);n=18581 | 1.83(1.78);n=17526 | <0.01 |
| 2.4(0.8);n=35391 | 2.39(0.81);n=18228 | 2.4(0.79);n=17163 | 0.01 |
| 1.2(0.3);n=36036 | 1.16(0.31);n=18544 | 1.18(0.31);n=17492 | 0.04 |
| 4.4(1.0);n=36150 | 4.3(1.0);n=18609 | 4.4(1.0);n=17541 | 0.02 |
| 9.2(3.5);n=33684 | 9.2(3.7);n=17561 | 9.1(3.4);n=16123 | 0.04 |
| 8.3(1.5);n=37166 | 8.3(1.6);n=19101 | 8.2(1.4);n=18065 | 0.06 |

After the propensity score matching, the baseline characteristic of the 2 groups were well-balanced, apart from eGFR with a difference of 1.6 between the 2 groups (SMD=0.13) (**Table 1**). The DPP4I and SGLT2I cohorts were comparable after matching with nearest neighbour search strategy with calliper of 0.1, and the proportional hazard assumption was confirmed **(Supplementary Figure 1)**. In the matched cohort, 197 patients developed new onset gastric cancer. Besides, 314 patients developed peptic ulcer, 246 developed acute gastritis, 325 developed non-acute gastritis, and 851 patients developed GERD. In total, 1960 patients passed away during the study period. The characteristics of patients are shown in **Table 1**.

### Association between SGLT2I and DPP4I and gastric cancer

In the matched cohort, 41 SGLT2I users and 156 DPP4I users developed gastric cancer. After a follow-up of 257947.7 person-year, the incidence of gastric cancer was lower amongst SGLT2I users (Incidence rate [IR] per 1000 person-year: 0.32; 95% CI: 0.23-0.43) compared to DPP4I users (IR per 1000 person-year: 1.22; 95% CI: 1.03-1.42) with a rate ratio of 0.26 (95% CI: 0.18-0.37) (**Table 2**). The incidence of gastric cancer amongst DPP4I users was similar to the incidence in China amongst patient over 60 years old (1.28 per 1000 person-time) [23, 24]. SGLT2I users had a 70% lower risk of gastric cancer after adjustment (HR: 0.30; 95% CI: 0.19-0.48, p<0.0001) compared to users of DPP4I regardless of the demographics, comorbidities, medication profile, renal function, inflammatory status, glycaemic tests, and the duration of T2DM (**Table 2; Supplementary Figure 2; Supplementary Table 3)**. This was substantiated by the cumulative incidence curves stratified by SGLT2I versus DPP4I (**Figure 2**).

**Table 2.** Incidence rate (IR) per 1000 person-year of primary outcomes and all-cause mortality in the SGLT2I v.s. DPP4I cohort before and after 1:1 propensity score matching. * for p≤ 0.05, ** for p ≤ 0.01, *** for p ≤ 0.001; HR: hazard ratio; CI: confidence interval; SGLT2I: sodium glucose cotransporter-2 inhibitor; DPP4I: dipeptidyl peptidase-4 inhibitor. Adjusted for significant demographics, past comorbidities, non-SGLT2I/DPP4I medications, abbreviated MDRD, NLR, fasting glucose, HbA1c, and duration from earliest diabetes mellitus date to initial drug exposure date.

| SGLT2i v.s. DPP4i |  |  |  |  |  |  |  |
| --- | --- | --- | --- | --- | --- | --- | --- |
| Gastric cancer |  |  |  | Peptic ulcer |  |  |  |
| DPP4i users | Person-year | Number of events | IR[95% CI] | DPP4i users | Person-year | Number of events | IR[95% CI] |
| Total | 128115.3 | 156 | 1.22[1.03-1.42] | Total | 127820.7 | 211 | 1.65[1.44-1.89] |
| SGLT2i users | Person-year | Number of events | IR[95% CI] | SGLT2i users | Person-year | Number of events | IR[95% CI] |
| Total | 129832.4 | 41 | 0.32[0.23-0.43] | Total | 129606.3 | 103 | 0.80[0.65-0.96] |
| Overall | 257947.7 | 197 | 0.76[0.66-0.88] | Overall | 257427 | 314 | 1.20[1.09-1.36] |
| Rate ratio |  |  | 0.26[0.18-0.37] | Rate ratio |  |  | 0.48[0.38-0.61] |
| Adjusted hazard ratio |  |  | 0.30[0.19-0.48]<br>P<0.0001*** | Adjusted hazard ratio |  |  | 0.66[0.47-0.91];<br>P=0.0118* |
| Acute gastritis |  |  |  | Non-acute gastritis |  |  |  |
| DPP4i users | Person-year | Number of events | IR[95% CI] | DPP4i users | Person-year | Number of events | IR[95% CI] |
| Total | 128061.7 | 185 | 1.44[1.24-1.67] | Total | 127828.5 | 246 | 1.92[1.69-2.18] |
| SGLT2i users | Person-year | Number of events | IR[95% CI] | SGLT2i users | Person-year | Number of events | IR[95% CI] |
| Total | 129749.4 | 61 | 0.47[0.36-0.60] | Total | 129718.7 | 79 | 0.61[0.48-0.76] |
| Overall | 257811.1 | 246 | 0.95[0.84-1.08] | Overall | 257547.2 | 325 | 1.25[1.21-1.30] |
| Rate ratio |  |  | 0.33[0.24-0.43] | Rate ratio |  |  | 0.31[0.25-0.41] |
| Adjusted hazard ratio |  |  | 0.31[0.20-0.48];<br>P<0.0001*** | Adjusted hazard ratio |  |  | 0.35[0.25-0.49];<br>P<0.0001*** |
| Gastroesophageal reflux disease |  |  |  | All-cause mortality |  |  |  |
| DPP4i users | Person-year | Number of events | IR[95% CI] | DPP4i users | Person-year | Number of events | IR[95% CI] |
| Total | 127052.7 | 495 | 3.90[3.56-4.25] | Total | 128619.7 | 1257 | 9.77[9.24-10.3] |
| SGLT2i users | Person-year | Number of events | IR[95% CI] | SGLT2i users | Person-year | Number of events | IR[95% CI] |
| Total | 128869.4 | 356 | 2.76[2.48-3.06] | Total | 129947.5 | 703 | 5.40[5.02-5.83] |
| **Overall** | 255922.1 | 851 | 3.32[3.11-3.56] | **Overall** | 258567.2 | 1960 | 7.58[7.25-7.92] |
| **Rate ratio** |  |  | 0.71[0.62-0.81] | **Rate ratio** |  |  | 0.55[0.50-0.61] |
| **Adjusted hazard ratio** |  |  | 0.62[0.50-0.76];<br>P<0.000*** | **Adjusted hazard ratio** |  |  | 0.77[0.66-0.89];<br>P=0.0003*** |

**Figure 2A.**
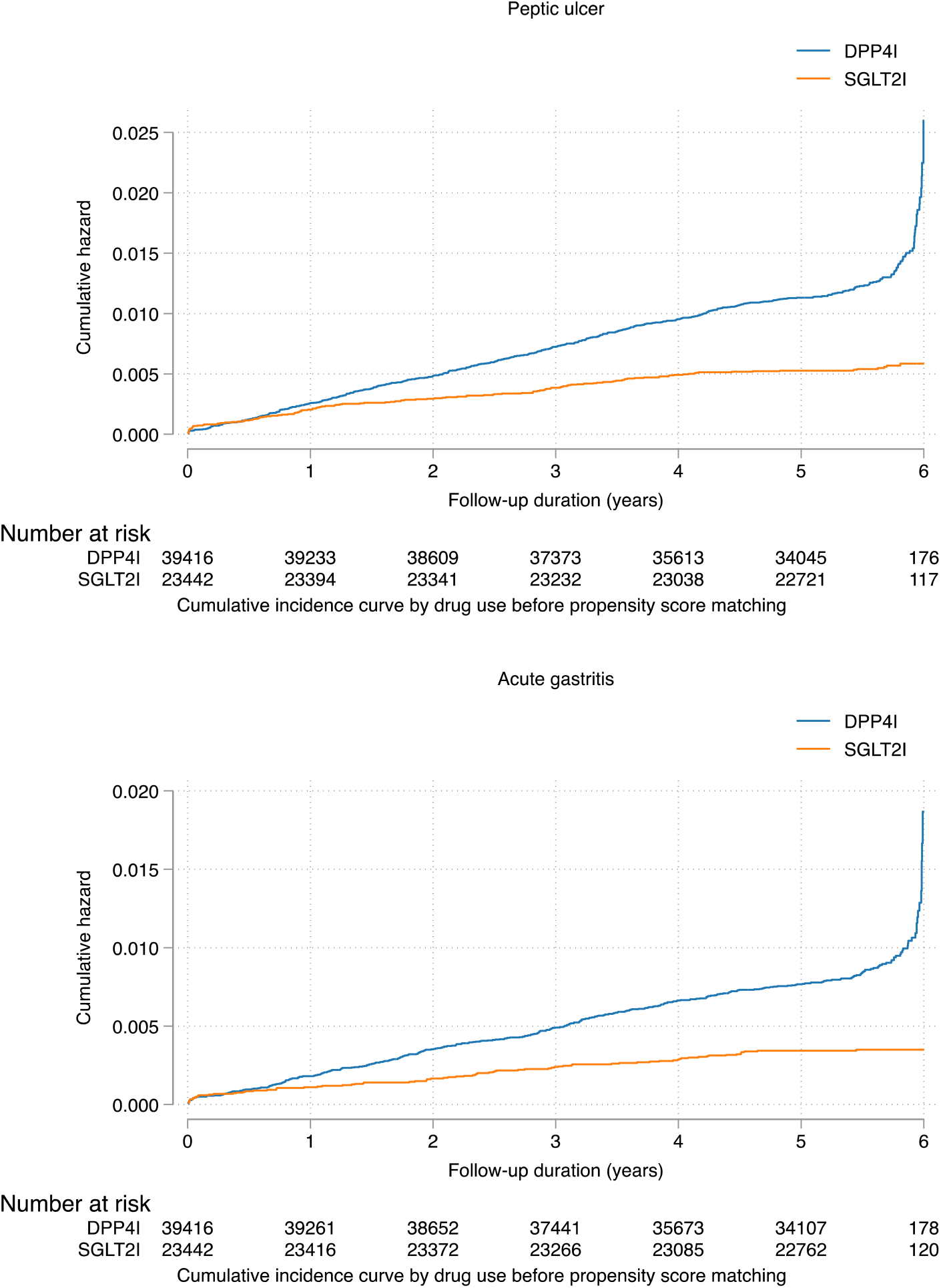
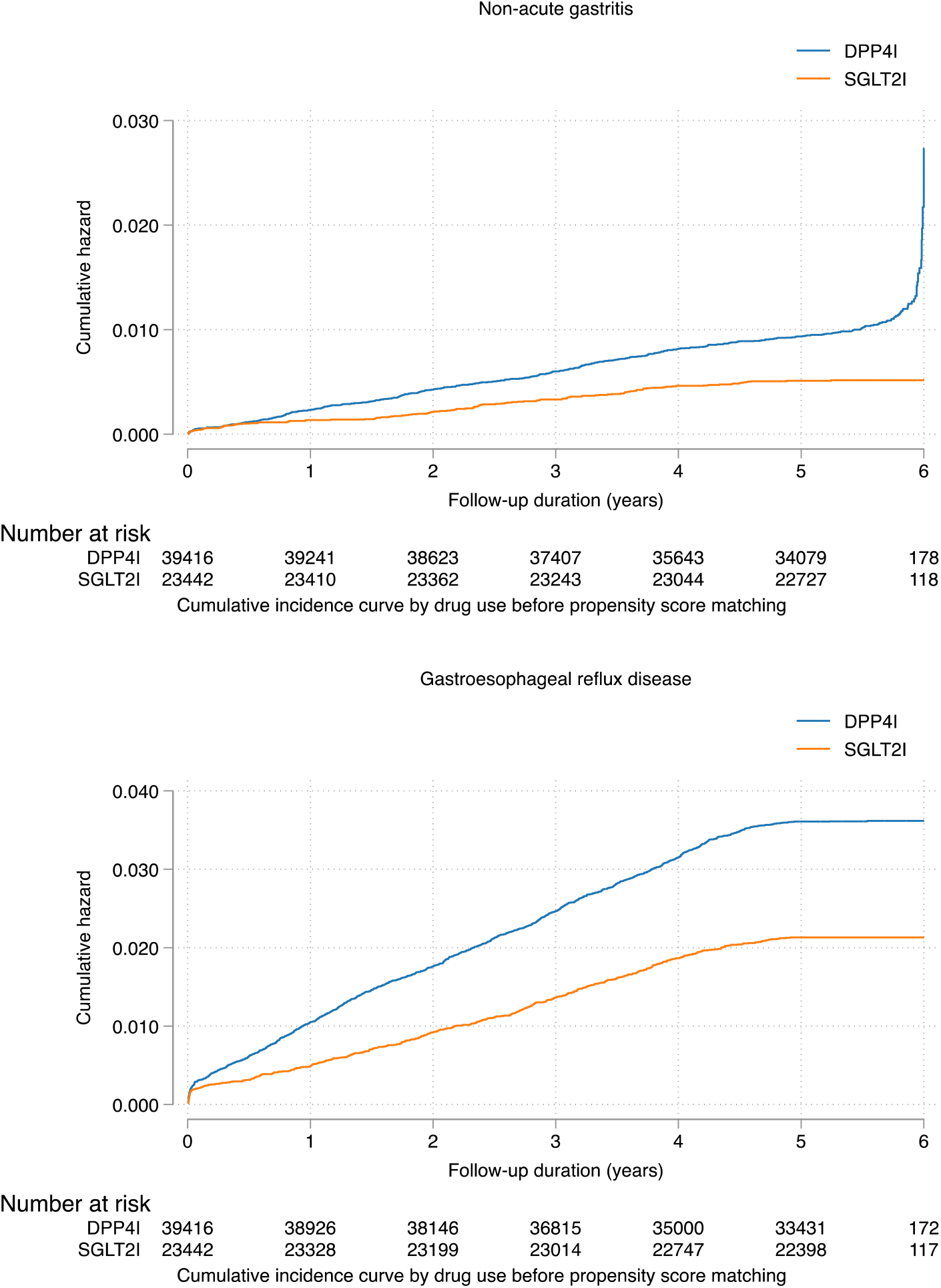
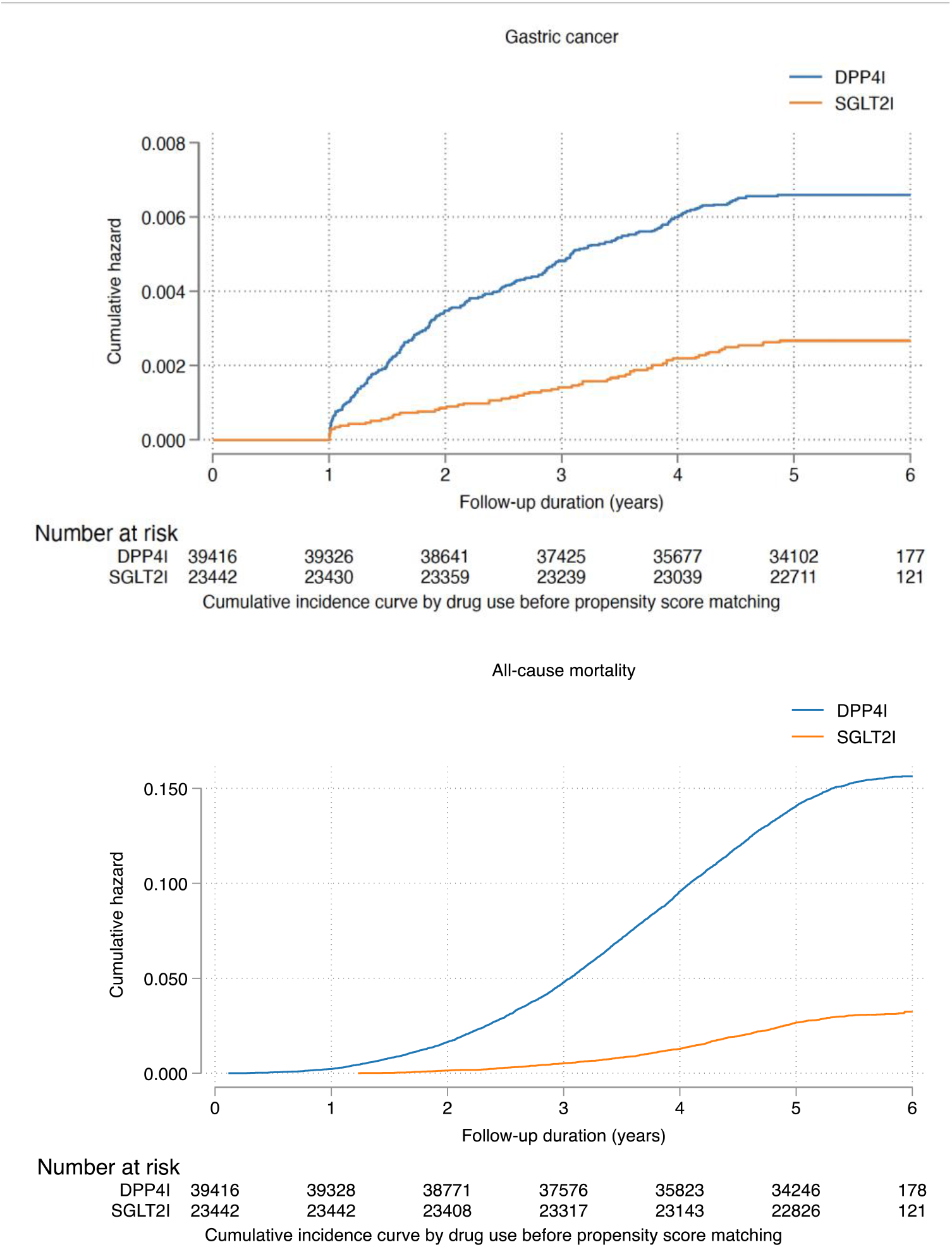
Cumulative incidence curves for new onset gastric outcomes and all-cause mortality stratified by drug exposure effects of SGLT2I and DPP4I before propensity score matching (1:1). SGLT2I: Sodium-glucose cotransporter-2 inhibitors; DPP4I: Dipeptidyl peptidase-4 inhibitors.

**Figure 2B.**
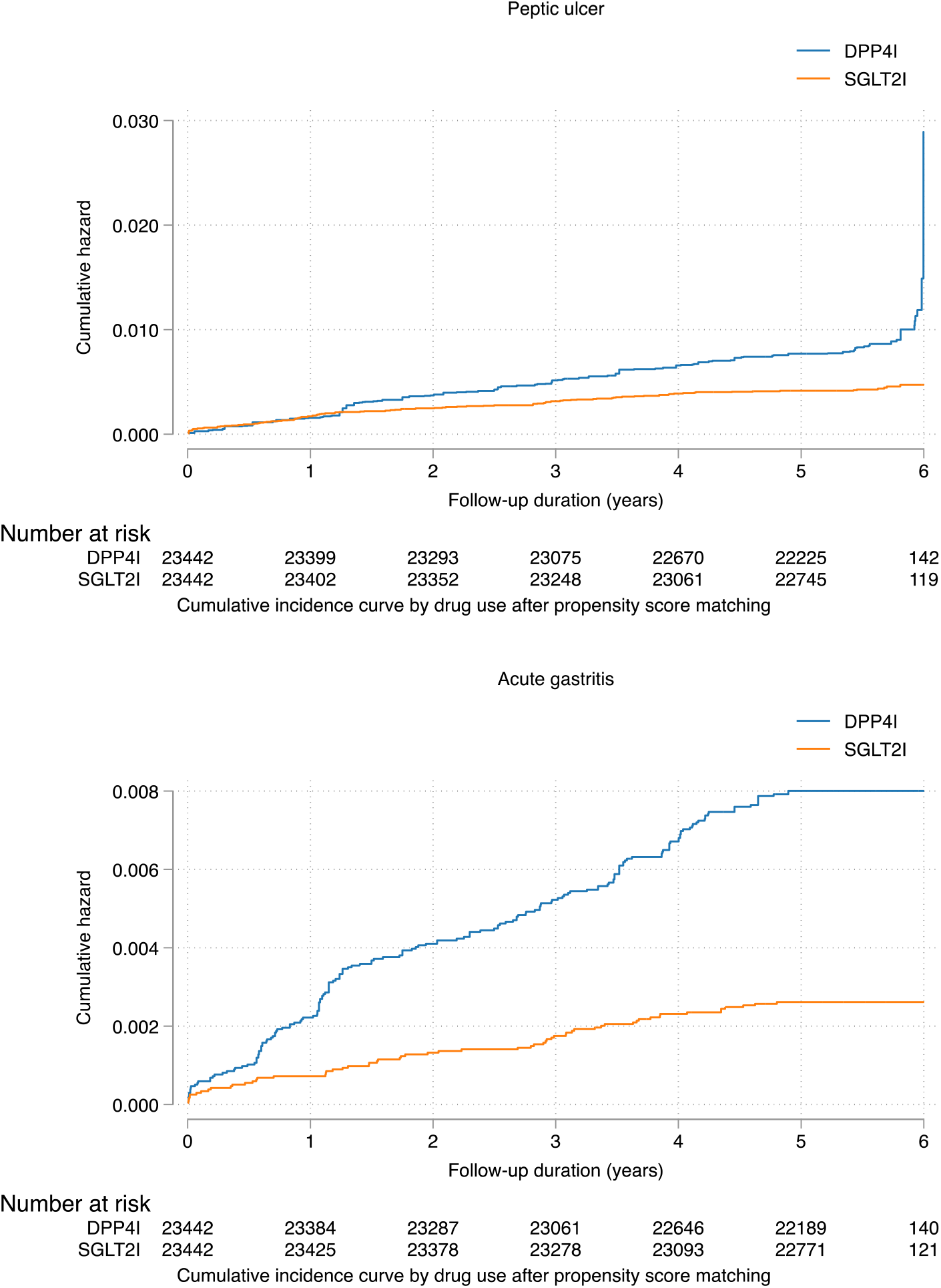
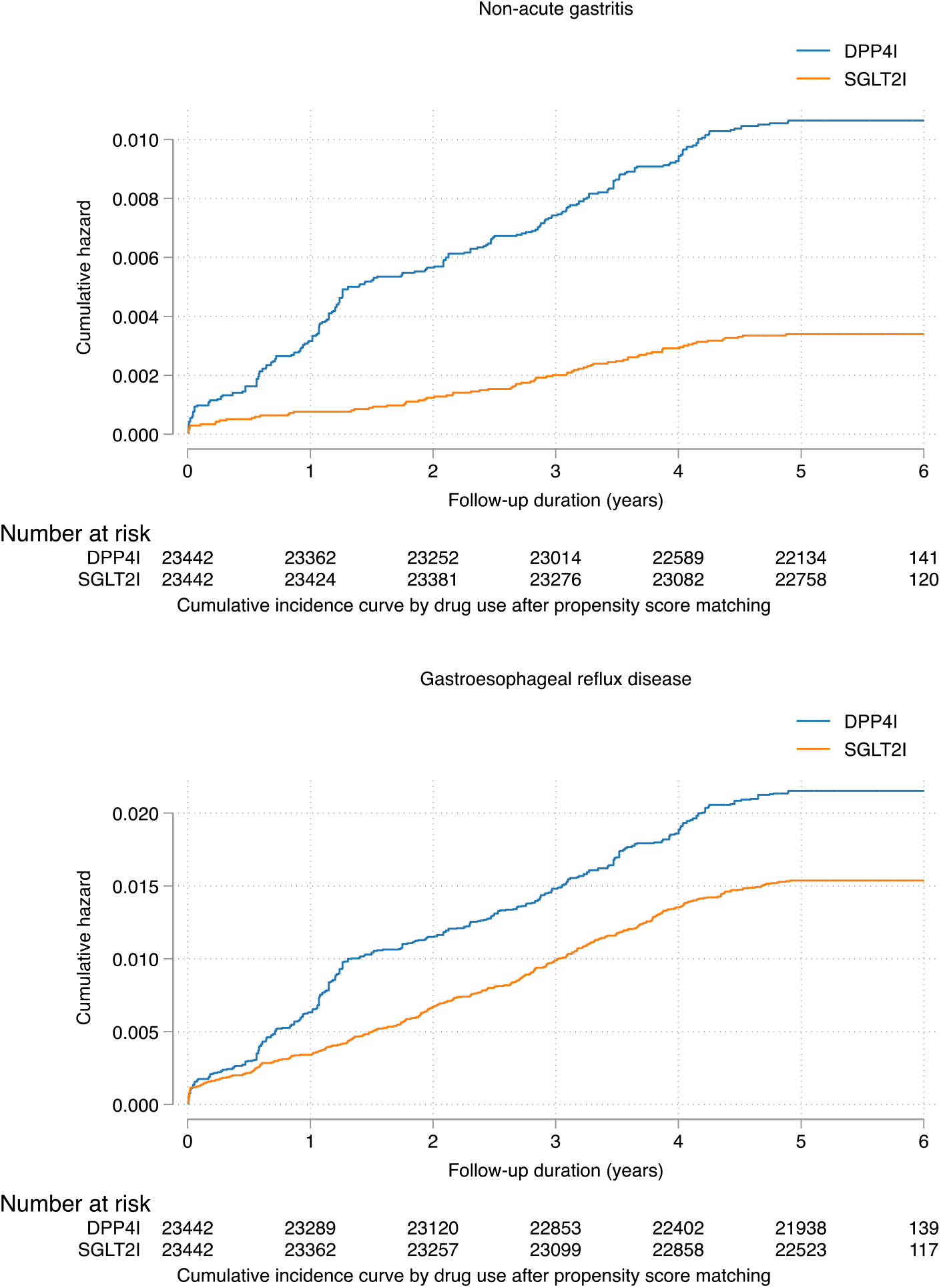
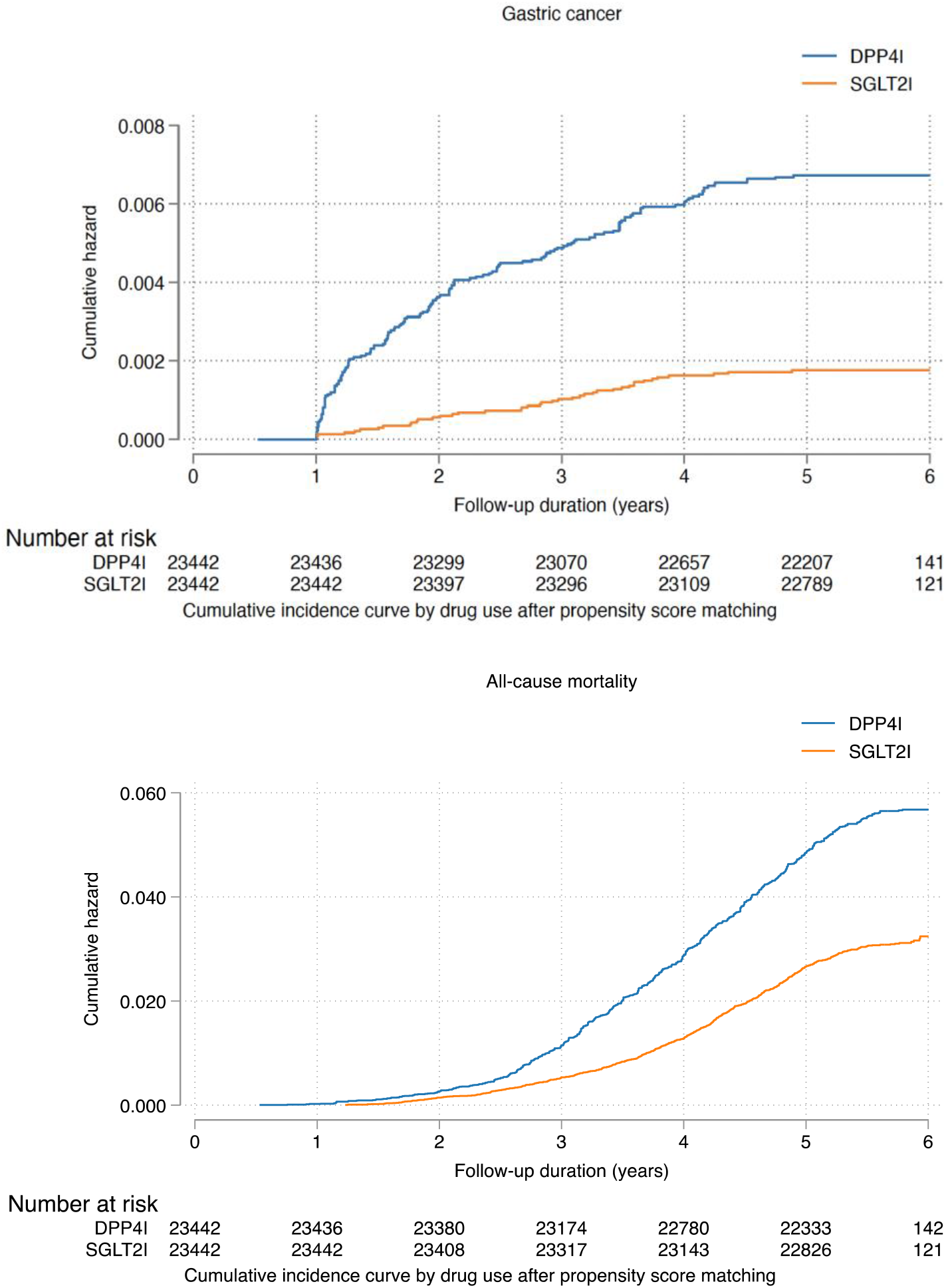
Cumulative incidence curves for new onset gastric outcomes and all-cause mortality stratified by drug exposure effects of SGLT2I and DPP4I after propensity score matching (1:1). SGLT2I: Sodium-glucose cotransporter-2 inhibitors; DPP4I: Dipeptidyl peptidase-4 inhibitors.

### Association between SGLT2I and DPP4I and gastric diseases

For peptic ulcer, 103 SGLT2I users and 211 DPP4I users developed peptic ulcer. After a total follow-up of 257427 person-year, the incidence of peptic ulcer was lower amongst SGLT2I users (IR per 1000 person-year: 0.80; 95% CI: 0.65-0.96) compared to DPP4I users (IR per 1000 person-year: 1.65; 95% CI: 1.44-1.89) with a rate ratio of 0.48 (95% CI: 0.38- 0.61) (**Table 2).** SGLT2I users had a 34% lower risk of peptic ulcer after adjustment (HR: 0.66; 95% CI: 0.47-0.91, p=0.0118) compared to DPP4I users (**Figure 2; Supplementary Figure 2; Supplementary Table 3).**

61 SGLT2I users and 185 DPP4I users developed acute gastritis. After a total follow-up of 257811.1 person-year, the incidence of acute gastritis was lower amongst SGLT2I users (IR per 1000 person-year: 0.47; 95% CI: 0.36-0.60) compared to DPP4I users (IR per 1000 person-year: 1.44; 95% CI: 1.24-1.67) with a rate ratio of 0.33 (95% CI: 0.24-0.43) (**Table 2).** SGLT2I users had a 69% lower risk of acute gastritis after adjustment (HR: 0.31; 95% CI: 0.20-0.48, p<0.0001) compared to DPP4I users (**Figure 2; Supplementary Figure 2; Supplementary Table 3)**.

For non-acute gastritis, 79 SGLT2I users and 246 DPP4I users developed non-acute gastritis. After a total follow-up of 257547.2 person-year, the incidence of non-acute gastritis was lower amongst SGLT2I users (IR per 1000 person-year: 0.61; 95% CI: 0.48-0.76) compared to DPP4I users (IR per 1000 person-year: 1.92; 95% CI: 1.69-2.18) with a rate ratio of 0.31 (95% CI: 0.25-0.41) (**Table 2).** SGLT2I users had a 65% lower risk of non-acute gastritis after adjustment (HR: 0.35; 95% CI: 0.25-0.49, p<0.0001) compared to DPP4I users (**Figure 2; Supplementary Figure 2; Supplementary Table 3)**.

For GERD, 356 SGLT2I users and 395 DPP4I users developed GERD. After a total follow-up of 255922.1 person-year, the incidence of GERD was lower amongst SGLT2I users (IR per 1000 person-year: 2.76; 95% CI: 2.48-3.06) compared to DPP4I users (IR per 1000 person-year: 3.90; 95% CI: 3.56-4.25) with a rate ratio of 0.71 (95% CI: 0.62-0.81) (**Table 2).** SGLT2I users had a 38% lower risk of GERD after adjustment (HR: 0.62; 95% CI: 0.50-0.76, p<0.0001) compared to DPP4I users (**Figure 2; Supplementary Figure 2; Supplementary Table 3)**.

### Secondary outcomes

703 SGLT2I users and 1257 DPP4I users passed away. After a follow-up of 258567.2 person-year, the incidence of all-cause mortality was lower amongst SGLT2I users (IR: 5.40; 95% CI: 5.02-5.83) compared to DPP4I users (IR: 9.77; 95% CI: 9.24-10.3) with a rate ratio of 0.26 (95% CI: 0.18-0.37) (**Table 2**). SGLT2I users had a 70% lower risk of all-cause mortality after adjustment (HR: 0.55; 95% CI: 0.50-0.61, p<0.0001) compared to users of DPP4I regardless of the duration of diabetes mellitus (**Supplementary Table 3)**. This was substantiated by the cumulative incidence curves stratified by SGLT2I versus DPP4I (**Figure 2; Supplementary Figure 2)**.

### Subgroup analysis

The results of the subgroup analysis for effects of SGLT2I and DPP4I on the gastric cancer and the gastric diseases are shown in **Figure 3**. For gastric cancer, the result demonstrated that SGLT2I was associated with lower risks of gastric cancer compared to DPP4I regardless of sex, age group, baseline HbA1c level, and medication uses. However, SGLT2I was not associated with lower risks of gastric cancer amongst patient with prior identified *H. pylori* infection and cancer **(Supplementary Figure 3 and 4**).

**Figure 3.**
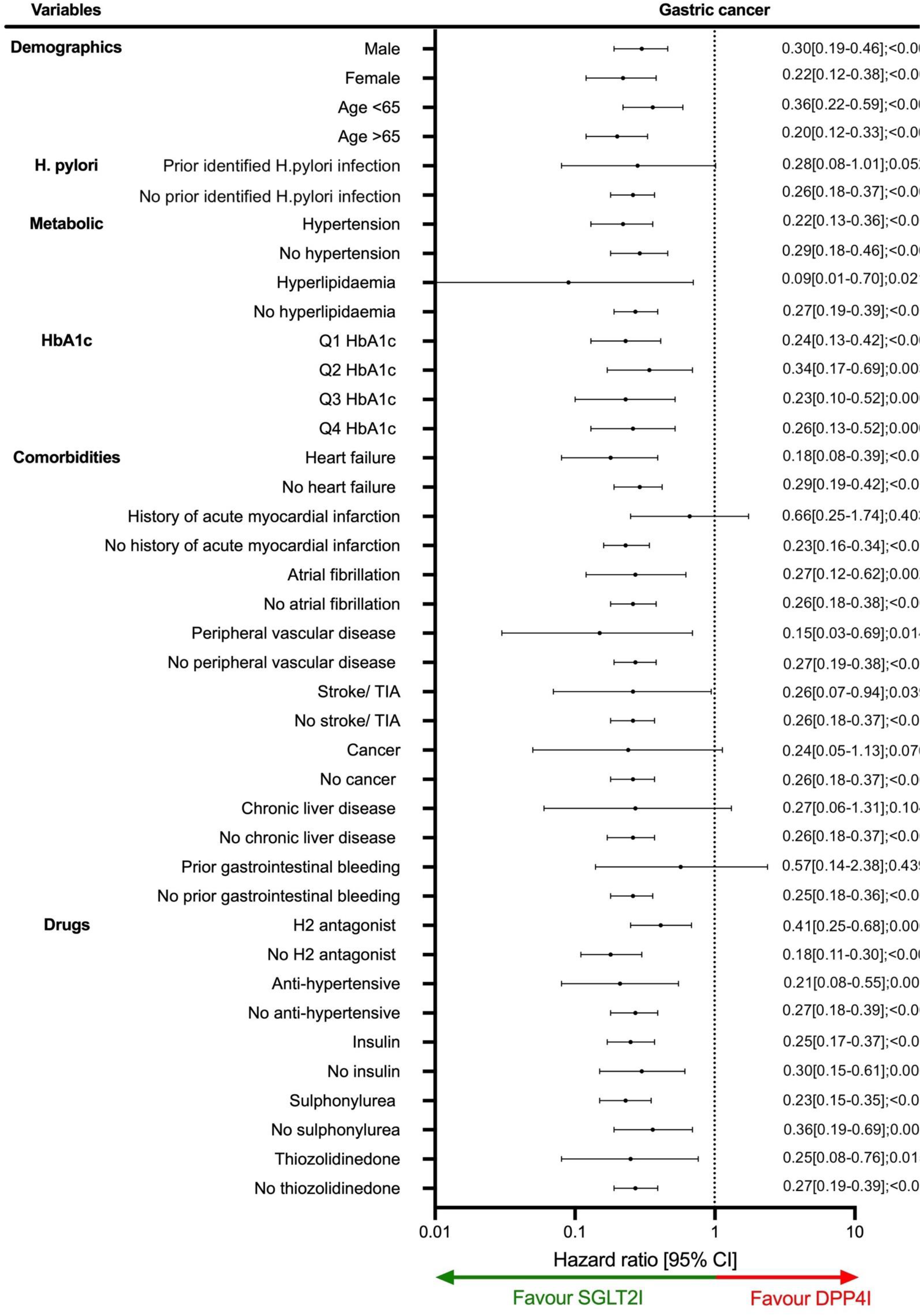
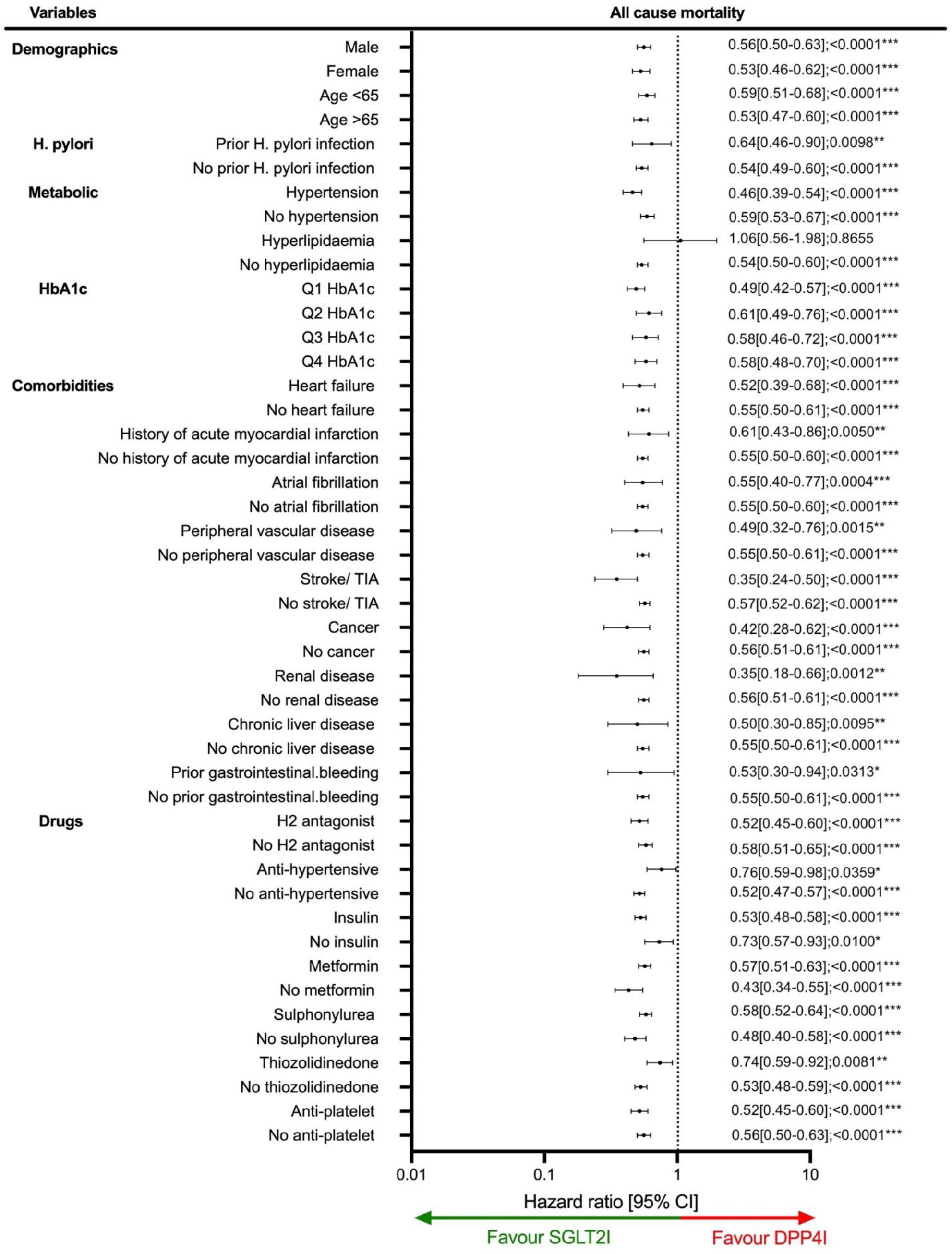
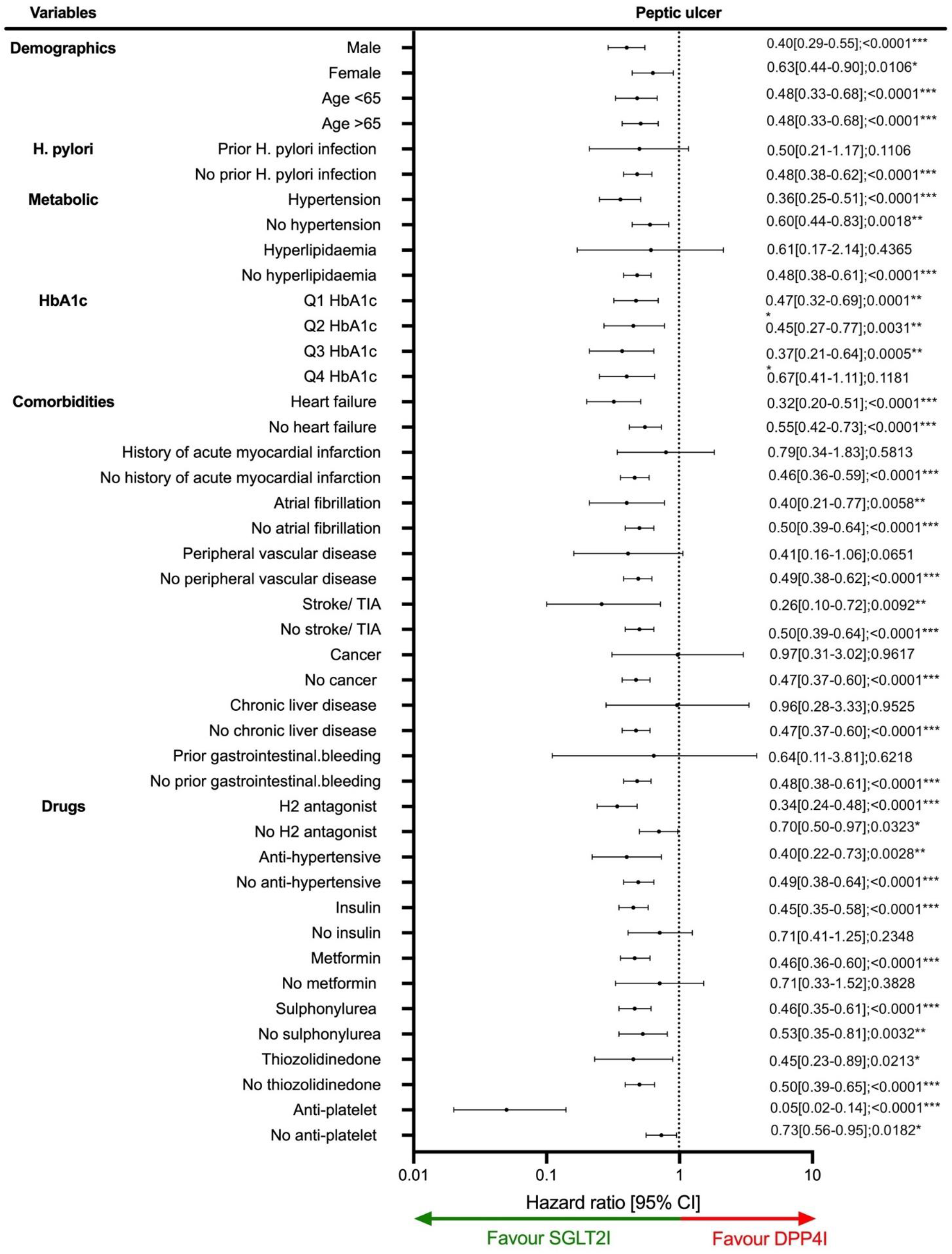
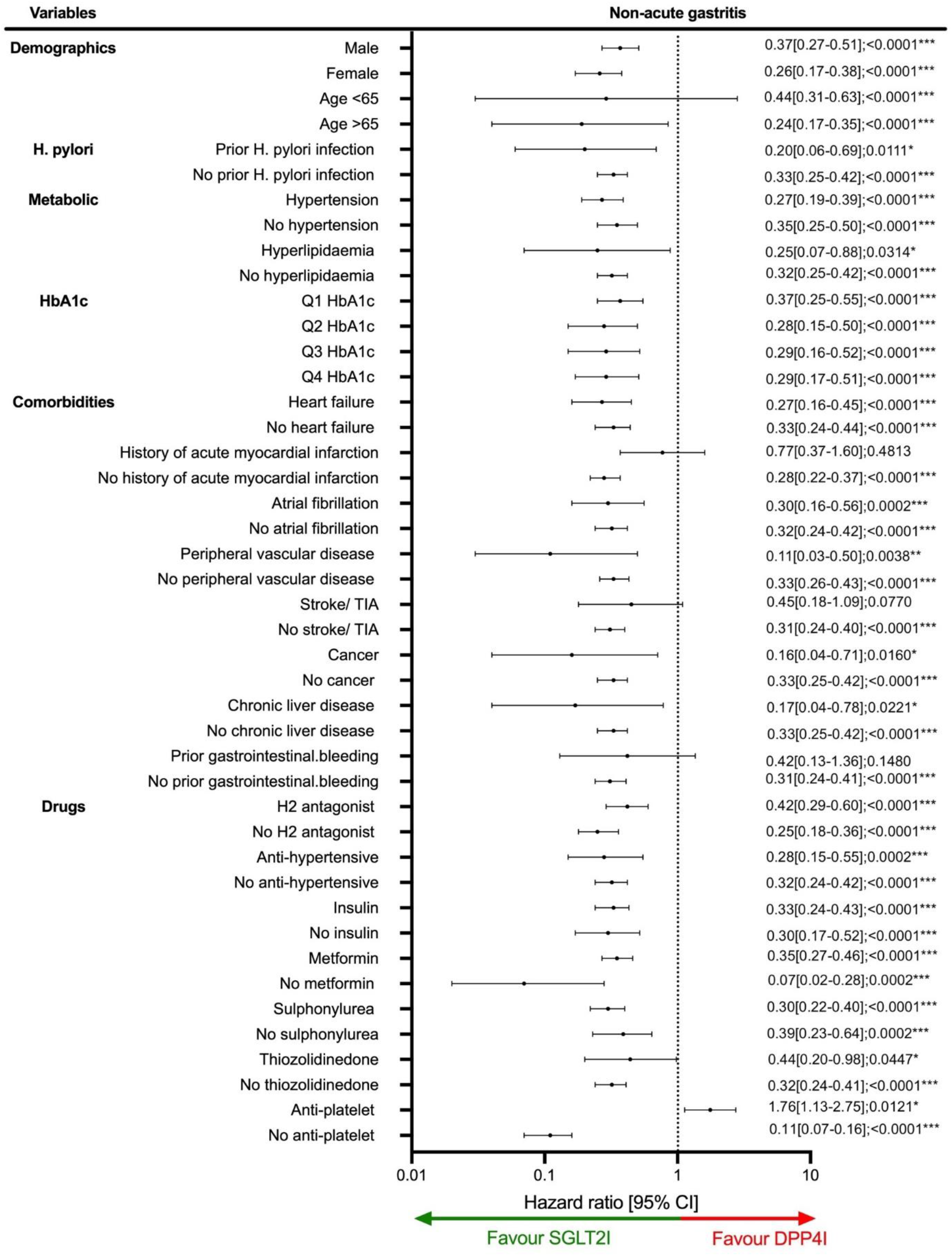
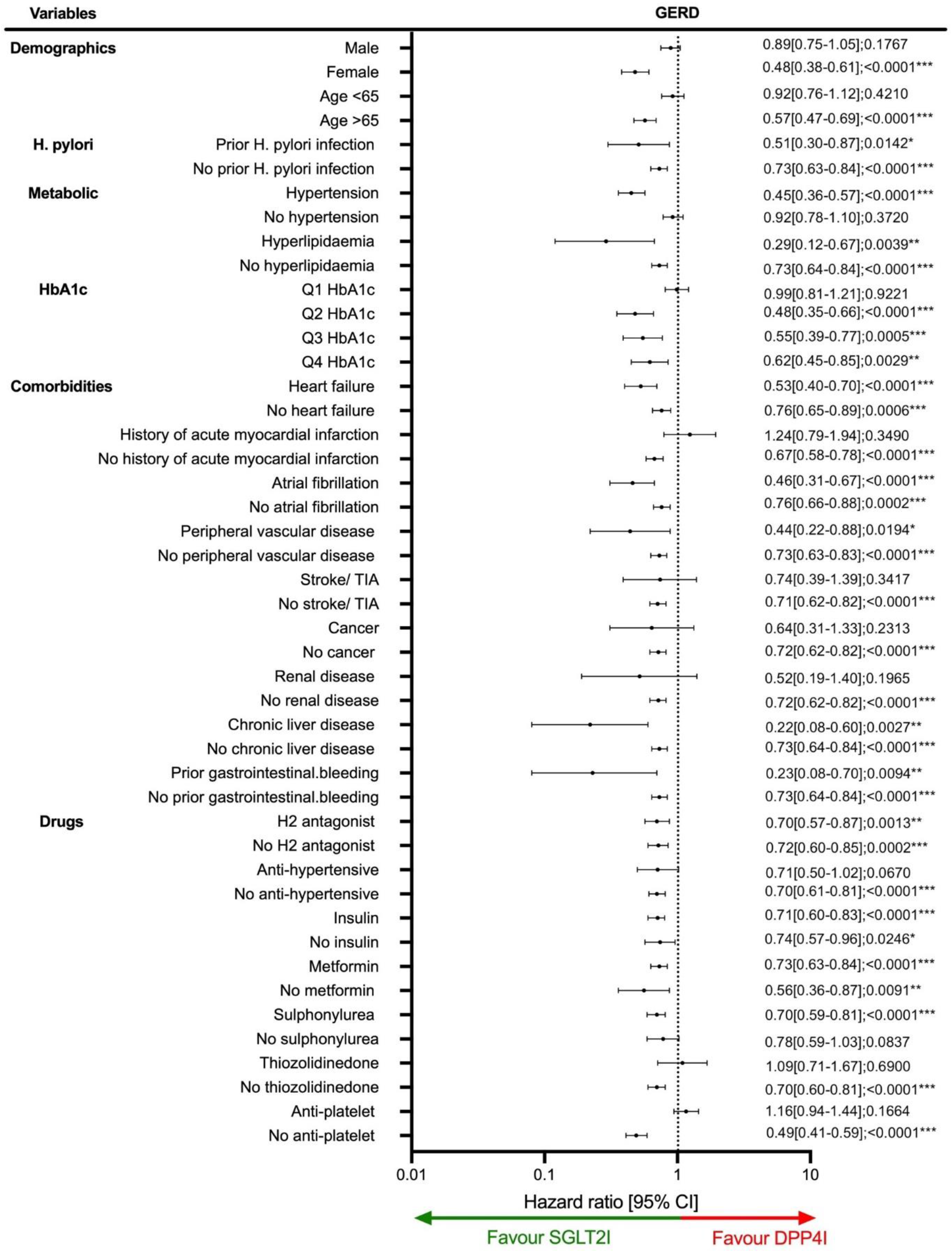
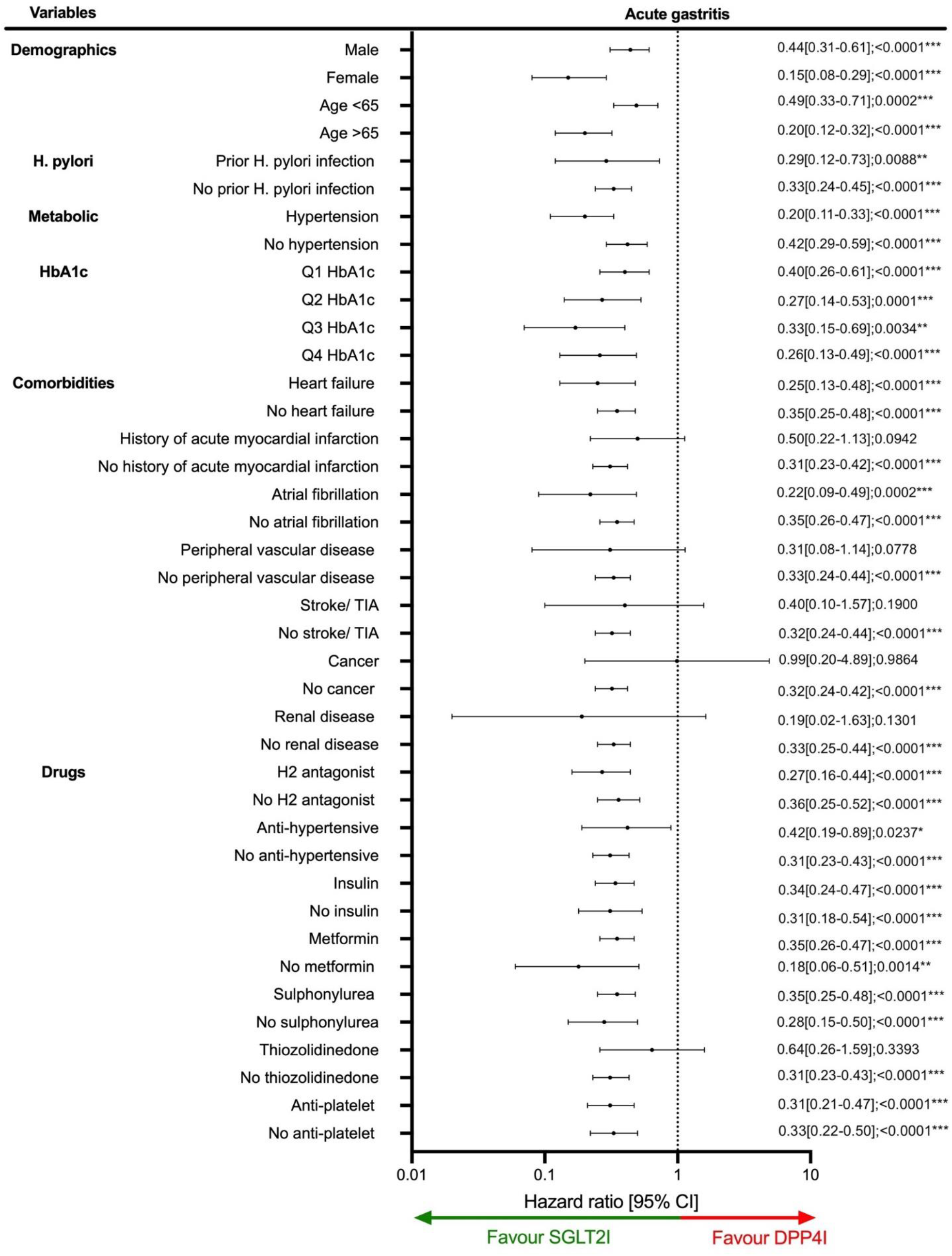
Subgroup analyses for SGLT2I v.s. DPP4I exposure predict new onset gastric cancer, gastric diseases, and all cause mortality in the matched cohort. SGLT2I: Sodium-glucose cotransporter-2 inhibitors; DPP4I: Dipeptidyl peptidase-4 inhibitors.Q1: Quartile 1Q2: Quartile 2; Q3: Quartile 3; Q4: Quartile 4; TIA: Transient ischaemic attack; CI: Confidence interval

For peptic ulcer, SGLT2I was associated with lower risks of peptic ulcer compared to DPP4I regardless of sex, age group, baseline HbA1c level and anti-platelet use. Similar to gastric cancer, amongst patient with prior identified *H. pylori* infection and cancer, SGLT2I was not associated with lower risks of peptic ulcer. For acute gastritis, SGLT2I was associated with lower risks of acute gastritis compared to DPP4I regardless of sex, age group, baseline HbA1c level, anti-platelet use and identified *H. pylori* infection status. Meanwhile for non-acute gastritis, SGLT2I was associated with lower risks of non-acute gastritis regardless of sex and baseline HbA1c level. Amongst patients age <65 years old there were no significant difference between SGLT2I and DPP4I on non-acute gastritis. Furthermore, amongst anti- platelet users DPP4I was associated with lower risks of non-acute gastritis compared to its counterpart. For GERD, SGLT2I was associated with lower risks of GERD compared to DPP4I amongst female, patients older than 65 years old, patient without anti-platelet use, and regardless of prior *H. pylori* identification status. SGLT2I was also associated with lower all-cause mortality compared to DPP4I users across all subgroups apart from hyperlipidaemic patients (**Figure 3**).

### Sensitivity analysis

Sensitivity analyses were performed to confirm the predictability of the models. A 3 arm analysis with the inclusion of GLP1A including patients only on (SGLT2I, DPP4I, and GLP1A) using stabilized IPTW was conducted (**Supplementary Table 4**). The results between DPP4I and SGLT2I remained consistent with the main result (all p<0.05) (**Table 2**). GLP1A had a 147% higher risk of gastric cancer (HR: 2.47; 95% CI: 1.27-4.81, p=0.0079) after adjustment compared to users of SGLT2I. The result also demonstrated that GLP1A had a higher risk of GERD (HR: 1.43; 95% CI: 1.04-1.97, p=0.0299), but not peptic ulcer (HR: 1.57; 95% CI: 0.80-3.05, p=0.1864), acute gastritis (HR: 1.03; 95% CI: 0.49-2.19, p=0.9343), and non-acute gastritis (HR: 1.51; 95% CI: 0.99-2.32, p=0.5818) after adjustment compared to users of SGLT2I. Furthermore, DPP4I was associated with higher risks of all-cause mortality (HR: 2.57; 95% CI: 2.36-2.79, p<0.0001), while GLP1A risks of all-cause mortality (HR: 0.84; 95% CI: 0.61-1.15; p=0.2659) was not significantly different from SGLT2I.

The results of the cause-specific hazard models, sub-distribution hazard models and different propensity score approaches demonstrated that different models did not change the point estimates for both the primary and the secondary outcomes (all p<0.05) **(Supplementary Table 5)**. Excluding patients with CKD stage 4/5 (eGFR <30), peritoneal dialysis or haemodialysis in the matched cohort likewise demonstrated that SGLT2I was associated with lower risks of all gastric outcomes compared to DPP4I **(Supplementary Table 6)**. Meanwhile, the sensitivity analysis for 1-year lag time also demonstrated the same trend (all p<0.05) **(Supplementary Table 7)**. Furthermore, in the as-treat approach which accounted for the effects of the drug-switching or discontinuation did not change the results (all p<0.05) **(Supplementary Table 8)**.

## Discussion

In this territory-wide prospectively recorded cohort study, we used real-world data to compare the relationship between SGLT2I versus DPP4I on gastric cancer and gastric diseases (peptic ulcer, acute gastritis, non-acute gastritis, and GERD). Our results demonstrated that SGLT2I was associated with an 70% lower risk of gastric cancer, 34% low risks of peptic ulcer, 69% lower risks of acute gastritis, 65% low risks of non-acute gastritis, and 38% of GERD than DPP4I users. To the best of our knowledge, this was the first cohort study to investigate the association between the novel anti-diabetic drugs with gastric cancer and the gastric diseases.

### Comparison with previous studies – gastric cancer

Overall, the incidence rate of gastric cancer in our study parallels the previous studies [23, 24]. Multiple population-based observational studies have illustrated the protective effects of metformin on gastric cancer [25]. On the other hand, studies investigating this association on newer anti-diabetic medications remain scarce.

In our study, the IR of the gastric cancer amongst DPP4I was 1.22 per 1000 person-time (95% CI: 1.03-1.42), which is similar to the incidence in China amongst patient over 60 years old (1.28 per 1000 person-time) (**Table 2**) [23, 24]. Besides, the risks of gastric cancer of DPP4I was similar to GLP1A in the sensitivity analysis. This suggested that DPP4I did not increase the risks of gastric cancer, but rather, SGLT2I might reduce the risks of gastric cancer.

The protective effects of SGLT2I and DPP4I against gastric cancer remain controversial as current literature demonstrates conflicting evidence. There have been some promising evidence revealing decreased risks of gastric cancer observed with canagliflozin use [10]. This is further supported by other meta-analysis studies which highlights the decreased risk of GI cancer in SGLT2I usage [26, 27]. Researchers hypothesize that the inhibitory effects of canagliflozin on SGLT1 and SGLT2 receptors can inhibit the glucose uptake of cancer cells, thereby increasing cell necrosis and reducing tumour growth [28].

A separate meta-analysis demonstrated that there was no significant increase in digestive system cancer risk observed in DPP4I usage compared to placebo (RR: 0.93 [0.77- 1.13]) [29]. This is further corroborated by another study that show DPP4I usage was not associated with increased gastric cancer risk when comparing DPP4I, GLP1A and metformin usage [30]. In our study, the sensitivity analysis result demonstrated that DPP4I and GLP1A shared similar risks of gastric cancer compared to SGLT2I **(Supplementary Table 4**). Wong *et al.* revealed that the addition of DPP4I for diabetic patients on metformin-sulfonylurea therapy correlated with the lowest risk of overall cancer compared to insulin and thiazolidinediones [31]. This may be attributed to a combination of the immunological function of DPP4 in activating quiescent T-lymphocytes and effects on secretion of cytotoxic granzymes, leading to cell apoptosis and decreasing carcinogenesis [32]. However, it must be noted that there are currently no studies investigated the direct relationship between DPP4I and gastric cancer in T2DM patients. Several studies examined the protective effects of GLP1A against prostate, breast and cervical cancer [33, 34, 35]. The GLP1A cohort data was utilised as a form of sensitive analysis to confirm their overall effects on mortality given its relatively small sample size compared to SGLT2I and DPP4I. As GLP1A is becoming more popular within the market in Hong Kong, the GLP1A cohort shall be expanded in few years’ time and its gastric effects can be further elucidated.

### Comparison with previous studies – gastric diseases

The incidence of peptic ulcers in this cohort is similar to the previous studies [36]. Current investigation surrounding the influence of SGLT2I, DPP4I and GLP1A on peptic ulcer is predominately conducted through animal models. Fujiwara *et al.* demonstrated that DPP4I inhibition using sitagliptin was effective against intestinal ulcers and improved ulcer healing through the activation of GLP-2 signal pathway [37]. This effect may be extrapolated to why anti-diabetic medications could generate a protective effect against peptic ulcer.

A meta-analysis demonstrated conflicting results regarding the effect of SGLT2I, DPP4I and GLP1A on various types of gastritis. In general, no significant association between SGLT2I and DPP4I and gastric diseases was found, whereas some GLP1A medications were associated with increased risks of some gastric diseases [38]. Furthermore, some studies elucidate that GLP1A usage correlated with increased gastritis and GERD [39]. The use of empagliflozin was associated with lower gastritis scores and milder inflammation presentation compared to the control group in rats [9].

SGLT2I and DPP4I agents have shown to facilitate improvement in GERD-like symptoms. Diabetic patients were linked to higher risks of GERD, as a result of autonomic neuropathy [40]. Compared to Western countries, GERD is relatively less common in Asia, which parallels the incidence rate presented in this study cohort [41]. An analysis using Japanese adverse drug event report database (JADER) highlighting DPP4Is usage showed a lower incidence of GERD-like symptoms compared to GLP1A usage [42]. In a randomized controlled trial, the use of lixisenatide and liraglutide revealed no significant influence on GERD or gastric motility [43].

### Clinical implications

The secondary protective benefits of SGLT2I and DPP4I on cardiovascular and gastric diseases have received worldwide attention in recent years. While there is more substantial evidence supporting the favourable effects of antidiabetic medications on cardiovascular mortality [44], there is limited evidence surrounding the latter. This notwithstanding, T2DM patients are susceptible to higher risk and mortality of gastric cancer [45, 46]. Therefore, the continuous elucidation of the possible implications of anti-diabetic medications is crucial for optimising management of gastric diseases, reducing healthcare resources and improving prognosis. In this study, we investigated the association using data from routine clinical practice, the result of which influence the choice of second-line antidiabetic therapy in T2DM patients regarding their gastric safety profile. As the results suggest, SGLT2I usage may contain a stronger protective effect against gastric cancer compared to DPP4I.

### Strengths and limitations

The strength of this study is the usage of CDARS, a comprehensive electronic health record database that includes extensive details of patient information such as drug prescription dates, time-serial laboratory results, and comorbidities. Hence, this limits information and selection and recall bias. The majority of current studies examine the effect of anti-diabetic medications as an individual predictor or comparing two of the drugs, whereas our study encompasses a comparison of two drugs with one extra drug (GLP1A) for sensitivity analysis. Furthermore, only new users of SGLT2I and DPP4I were included in this study, such that the effects of the baseline drug will be minimised.

In contrast, several limitations should be acknowledged. First, due to the observational nature of this study, common variables including smoking, drinking, BMI, and socioeconomic status were also not available from CDARS. This could only be addressed by including the comorbidities and the laboratory tests results in the study, to indirectly refer their cardiovascular risk factors. However, since the cohorts were well-matched over wide range of diseases and medications, ideally the covariates not included should be well-balanced. The data results may be susceptible to coding errors, missing data, under-coding and underreporting of clinical diagnoses. To minimise this, we have included all available data on laboratory tests and medications, as well as verifying the ICD-9 diagnosis codes with existing studies. As the accurate medications and laboratory testing results were matched and also adjusted in the regression, the impacts of the coding error effects may be compensated.

Besides, the drug exposure duration, patient compliance to medication could not be standardised. The patient’s level of medication adherence was indirectly assessed through the frequency prescription refills. This may lead to time lag biases and immortal time. Additionally, patients who may have switched between SGLT2I or DPP4I usage due to the presence of comorbidities or poor glycaemic control, may have contributed to worse gastric outcomes among SGLT2I users. Nonetheless, this would result in an underestimation of the protective influences of SGLT2I against gastric outcomes and would not have a drastic impact on the overall results.

Nevertheless, it was a common practice to get the ICD-9 code for patients with *H. pylori* infection diagnosed after oesophagogastroduodenoscopy (OGD), hence it is possible that there may be an underestimation of *H. pylori* in ICD9 as not all patients may undergo OGD and tissue sampling. Further research is warranted to explore the effects of serological gastritis markers and endoscopic findings on gastric outcomes. Lastly, the nature of the study design suggests that the findings between specific drugs and gastric outcomes are correlational in nature. Thus, prospective randomised controlled trials are imperative to evaluate the causal links of anti-diabetic medications.

## Conclusions

In this population-based cohort study, SGLT2I users was associated with lower risks of new-onset gastric cancer compared to DPP4I users. SGLT2I users was also associated with lower risks of peptic ulcer, acute gastritis, non-acute gastritis, and GERD after matching and adjustments compared to DPP4I users. These results may have potential clinical implications in reducing the gastric complications. Further mechanistic study into the gastric cancer with SGLT2I are needed.

## Funding source

The authors received no funding for the research, authorship, and/or publication of this article.

## Conflicts of Interest

None.

## Availability of data and materials

An anonymised version without identifiable or personal information is available from the corresponding authors upon reasonable request for research purposes.

## Data Availability

All data produced in the present study are available upon reasonable request to the authors

## Acknowledgements

None.

## Guarantor Statement

All authors approved the final version of the manuscript. GT is the guarantor of this work and, as such, had full access to all the data in the study and takes responsibility for the integrity of the data and the accuracy of the data analysis.

## Author contributions

Data analysis: OHIC, VKC, LL, JZ Data review: OHIC, SL, GT, JZ Data acquisition: OHIC, TTLL, SL

Data interpretation: OHIC, CTC, WTW, BMYC, GT, JZ Critical revision of manuscript: AK, WTW, BMYC, GT, JZ Supervision: BMYC, WTW, GT, JZ

Manuscript writing: OHIC, CTC, ZMWN, KKW

Manuscript revision: OHIC, TTLL, ZMWN, KKW, SL, HL, TKRP

## Notes

### Competing Interest Statement

The authors have declared no competing interest.

### Funding Statement

This study did not receive any funding

### Author Declarations

This study was approved by the Institutional Review Board of the University of Hong Kong/Hospital Authority Hong Kong West Cluster and The Joint Chinese University of Hong Kong-New Territories East Cluster.

