## Supplementary Appendix for "Comparing the risks of new-onset gastric cancer or gastric diseases in type 2 diabetes mellitus patients exposed to SGLT2I, DPP4I or GLP1A: a population-based cohort study"

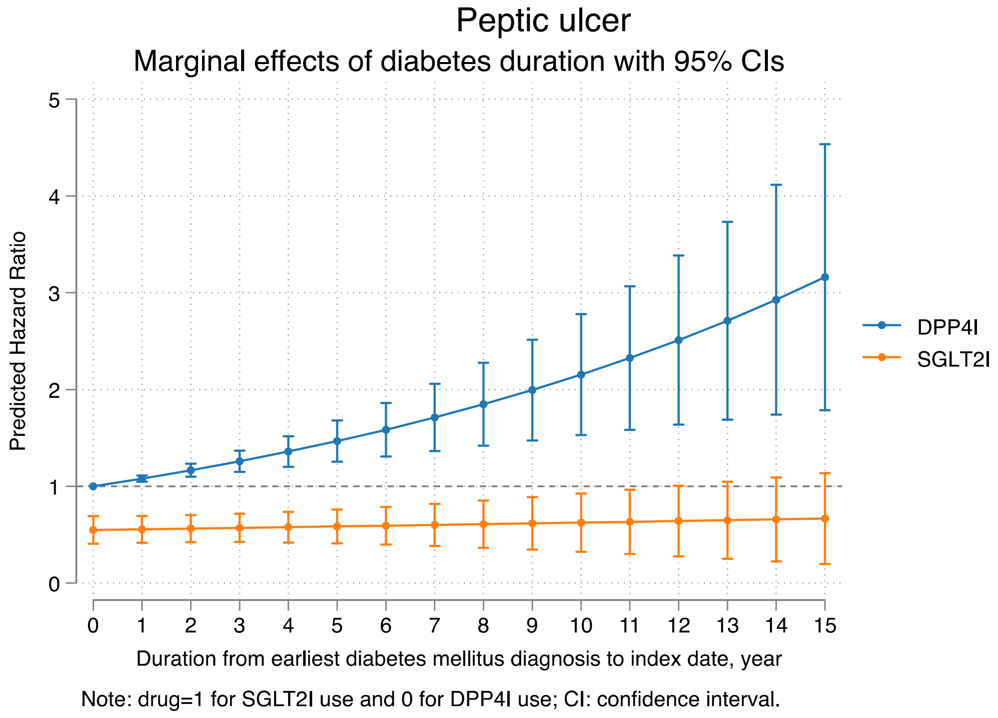
Peptic ulcer
Marginal effects of diabetes duration with 95% CIs
Predicted Hazard Ratio
0
1
2
3
4
5
0 1 2 3 4 5 6 7 8 9 10 11 12 13 14 15
Duration from earliest diabetes mellitus diagnosis to index date, year
DPP4I
SGLT2I
Note: drug=1 for SGLT2I use and 0 for DPP4I use; CI: confidence interval.

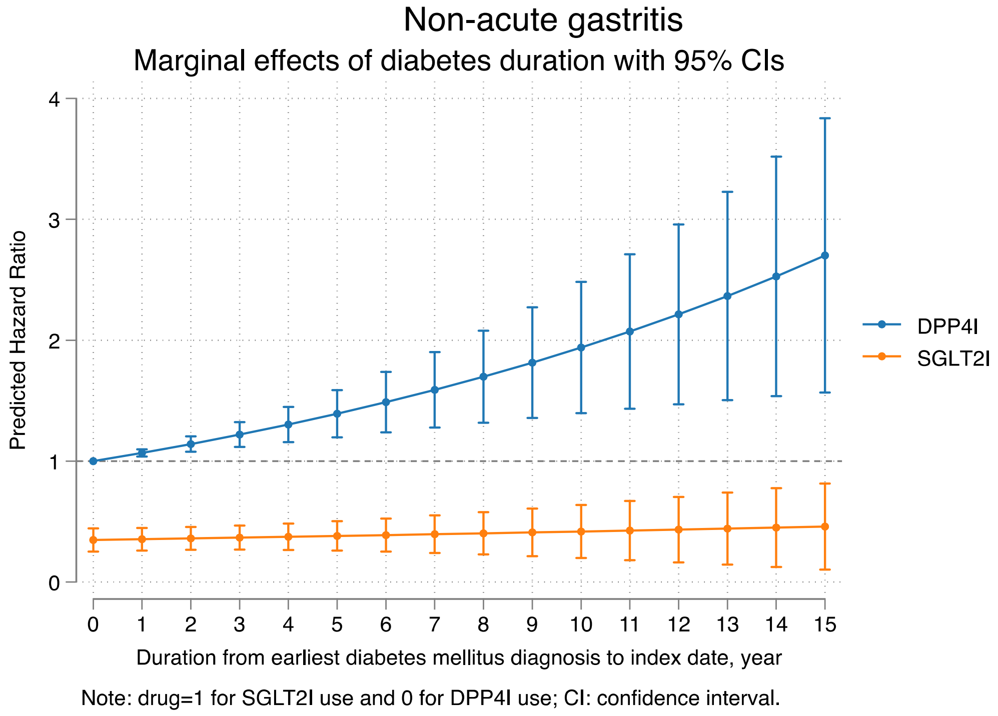
Non-acute gastritis
Marginal effects of diabetes duration with 95% CIs
Predicted Hazard Ratio
0
1
2
3
4
0 1 2 3 4 5 6 7 8 9 10 11 12 13 14 15
Duration from earliest diabetes mellitus diagnosis to index date, year
DPP4I
SGLT2I
Note: drug=1 for SGLT2I use and 0 for DPP4I use; CI: confidence interval.

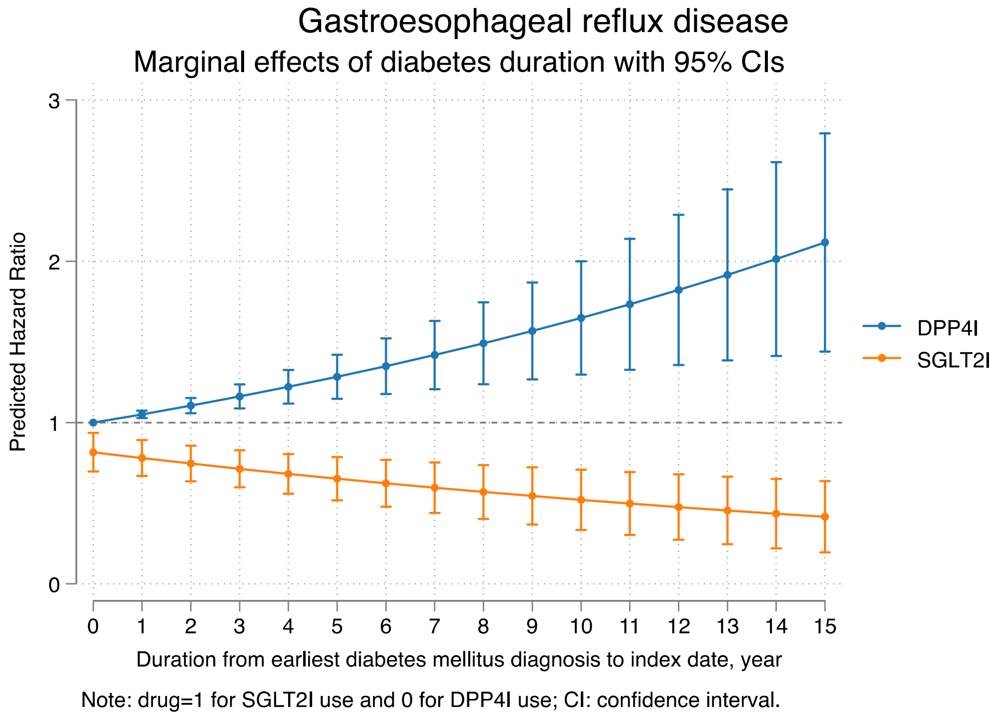
Gastroesophageal reflux disease
Marginal effects of diabetes duration with 95% CIs
Predicted Hazard Ratio
0
1
2
3
0 1 2 3 4 5 6 7 8 9 10 11 12 13 14 15
Duration from earliest diabetes mellitus diagnosis to index date, year
DPP4I
SGLT2I
Note: drug=1 for SGLT2I use and 0 for DPP4I use; CI: confidence interval.

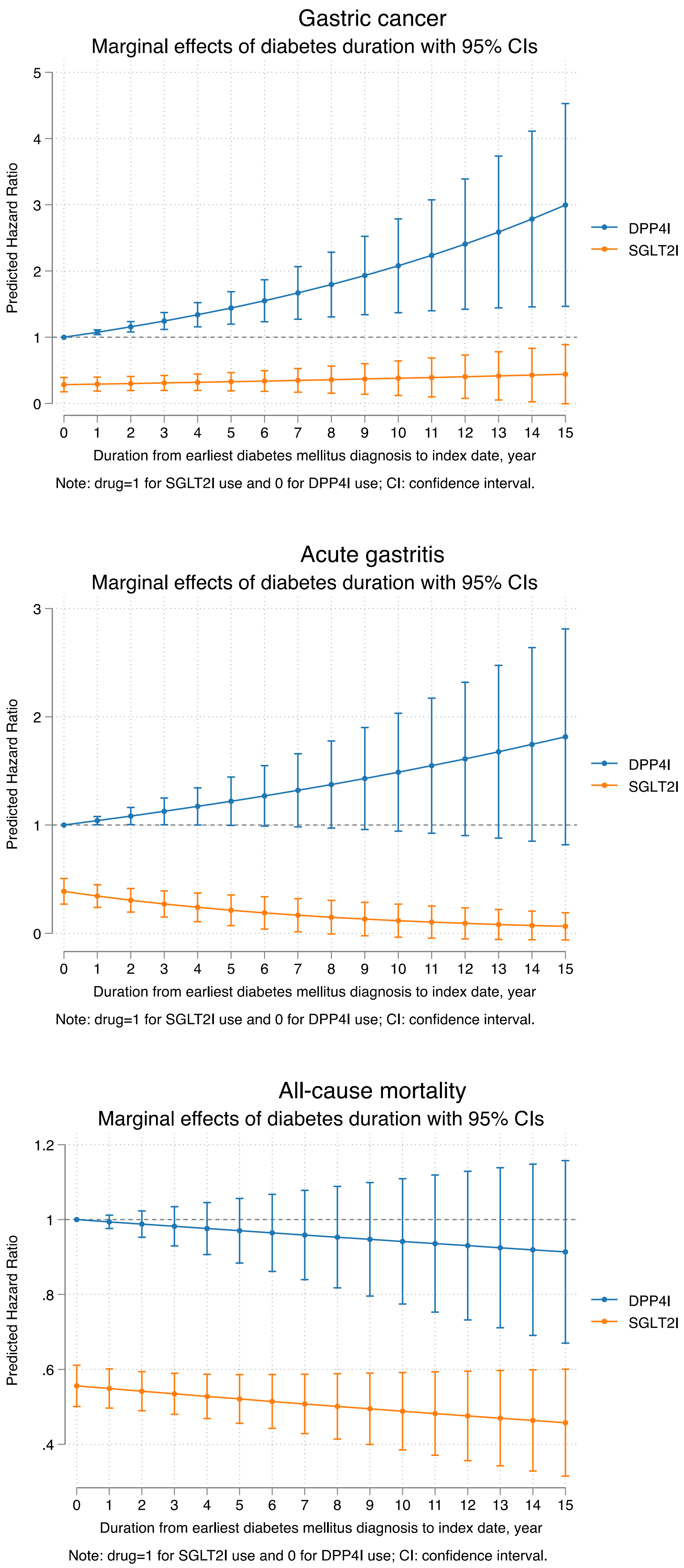

**Supplementary Figure 2. Marginal effects of diabetes duration with 95% CIs on new onset adverse gastric events and all-cause mortality stratified by drug use.**

SGLT2I: Sodium-glucose cotransporter-2 inhibitors; DPP4I: Dipeptidyl peptidase-4 inhibitors.

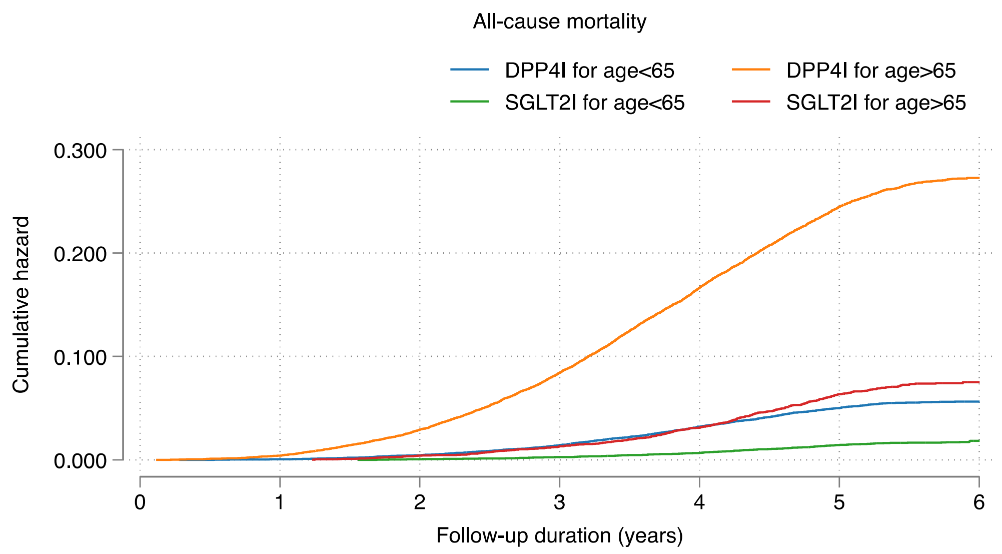

Number at risk

| | | | | | | | |
|---|---|---|---|---|---|---|---|
| DPP4I for age<65 | 20124 | 20113 | 20035 | 19841 | 19487 | 19142 | 101 |
| DPP4I for age>65 | 19292 | 19215 | 18736 | 17735 | 16336 | 15104 | 77 |
| SGLT2I for age<65 | 17489 | 17489 | 17479 | 17442 | 17373 | 17239 | 85 |
| SGLT2I for age>65 | 5953 | 5953 | 5929 | 5875 | 5770 | 5587 | 36 |

Cumulative incidence curve by drug use before propensity score matching

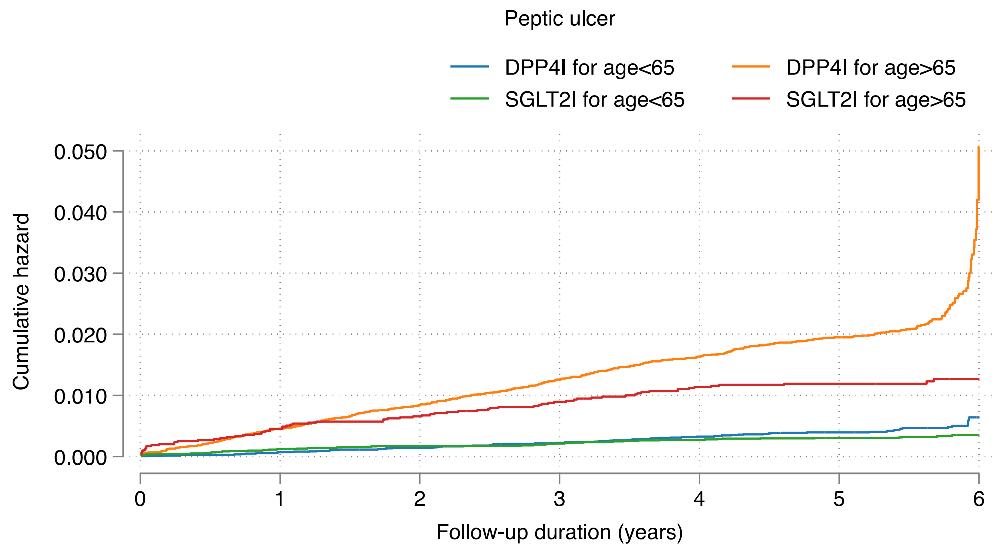

Number at risk

| | | | | | | | |
|---|---|---|---|---|---|---|---|
| DPP4I for age<65 | 20124 | 20099 | 20009 | 19804 | 19441 | 19094 | 101 |
| DPP4I for age>65 | 19292 | 19134 | 18600 | 17569 | 16172 | 14951 | 75 |
| SGLT2I for age<65 | 17489 | 17468 | 17449 | 17407 | 17327 | 17188 | 85 |
| SGLT2I for age>65 | 5953 | 5926 | 5892 | 5825 | 5711 | 5533 | 32 |

Cumulative incidence curve by drug use before propensity score matching

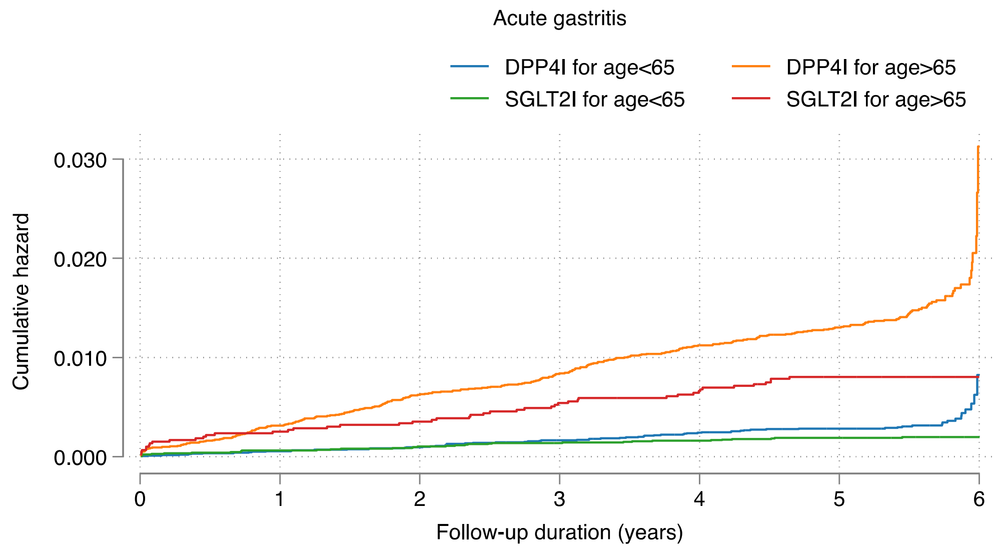

Number at risk

| | | | | | | | |
|---|---|---|---|---|---|---|---|
| DPP4I for age<65 | 20124 | 20102 | 20016 | 19811 | 19450 | 19107 | 101 |
| DPP4I for age>65 | 19292 | 19159 | 18636 | 17630 | 16223 | 15000 | 77 |
| SGLT2I for age<65 | 17489 | 17478 | 17462 | 17419 | 17347 | 17211 | 85 |
| SGLT2I for age>65 | 5953 | 5938 | 5910 | 5847 | 5738 | 5551 | 35 |

Cumulative incidence curve by drug use before propensity score matching

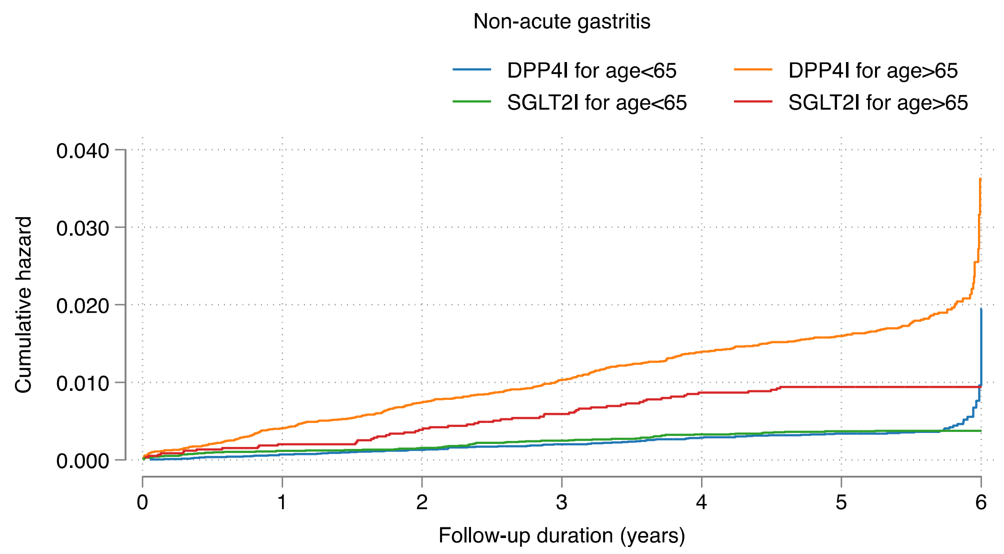

Number at risk

| | | | | | | | |
|---|---|---|---|---|---|---|---|
| DPP4I for age<65 | 20124 | 20099 | 20009 | 19805 | 19444 | 19097 | 101 |
| DPP4I for age>65 | 19292 | 19142 | 18614 | 17602 | 16199 | 14982 | 77 |
| SGLT2I for age<65 | 17489 | 17469 | 17452 | 17399 | 17317 | 17181 | 85 |
| SGLT2I for age>65 | 5953 | 5941 | 5910 | 5844 | 5727 | 5546 | 33 |

Cumulative incidence curve by drug use before propensity score matching

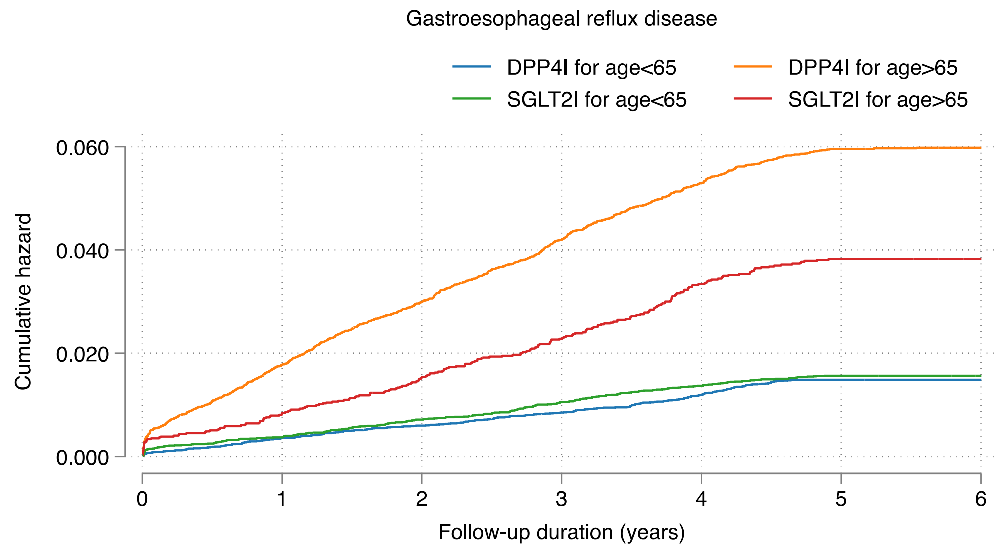

Number at risk

| | | | | | | | |
|---|---|---|---|---|---|---|---|
| DPP4I for age<65 | 20124 | 20041 | 19919 | 19689 | 19293 | 18919 | 100 |
| DPP4I for age>65 | 19292 | 18885 | 18227 | 17126 | 15707 | 14512 | 72 |
| SGLT2I for age<65 | 17489 | 17424 | 17357 | 17264 | 17147 | 16989 | 83 |
| SGLT2I for age>65 | 5953 | 5904 | 5842 | 5750 | 5600 | 5409 | 34 |

Cumulative incidence curve by drug use before propensity score matching

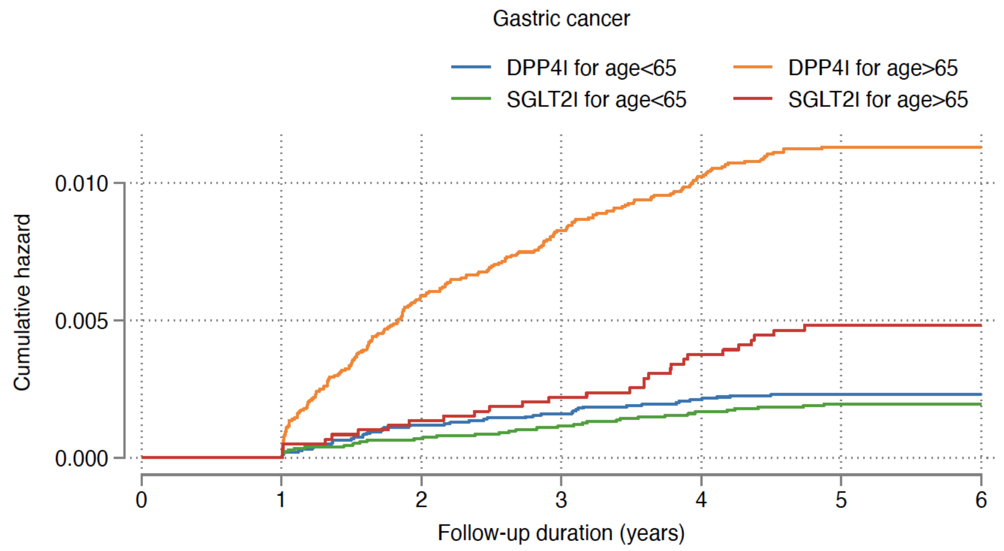

Number at risk

| | | | | | | | |
|---|---|---|---|---|---|---|---|
| DPP4I for age<65 | 20124 | 20113 | 20011 | 19812 | 19453 | 19107 | 101 |
| DPP4I for age>65 | 19292 | 19213 | 18630 | 17613 | 16224 | 14995 | 76 |
| SGLT2I for age<65 | 17489 | 17479 | 17446 | 17391 | 17309 | 17168 | 85 |
| SGLT2I for age>65 | 5953 | 5951 | 5913 | 5848 | 5730 | 5543 | 36 |

Cumulative incidence curve by drug use before propensity score matching

**Supplementary Figure 3A. Cumulative incidence curves for new onset gastric outcomes and all-cause mortality stratified by combinations of age and drug exposure effects of SGLT2I and DPP4I before propensity score matching.**
SGLT2I: Sodium-glucose cotransporter-2 inhibitors; DPP4I: Dipeptidyl peptidase-4 inhibitors.

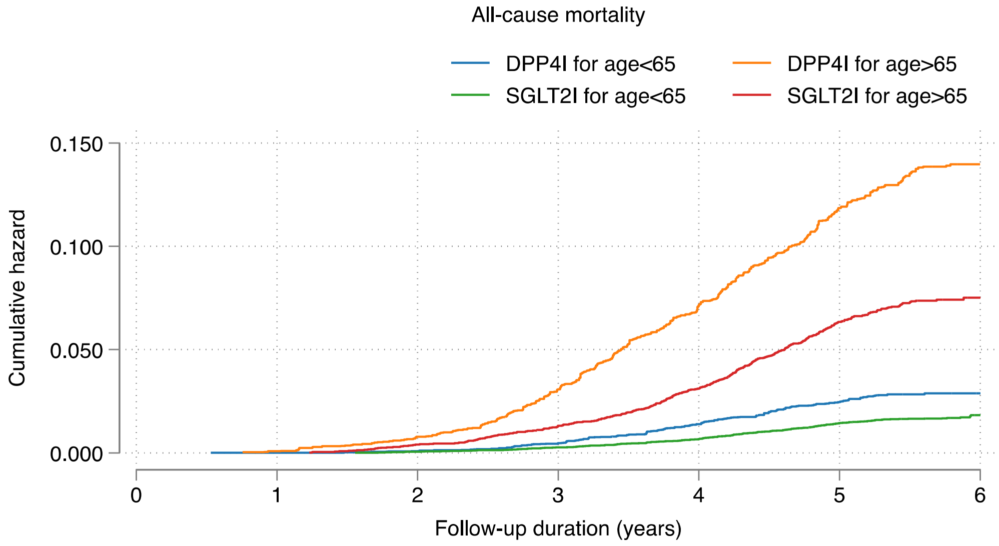

Number at risk

| | 0 | 1 | 2 | 3 | 4 | 5 | 6 |
|---|---|---|---|---|---|---|---|
| DPP4I for age<65 | 17269 | 17268 | 17255 | 17188 | 17030 | 16850 | 90 |
| DPP4I for age>65 | 6173 | 6168 | 6125 | 5986 | 5750 | 5483 | 52 |
| SGLT2I for age<65 | 17489 | 17489 | 17479 | 17442 | 17373 | 17239 | 85 |
| SGLT2I for age>65 | 5953 | 5953 | 5929 | 5875 | 5770 | 5587 | 36 |

Cumulative incidence curve by drug use after propensity score matching

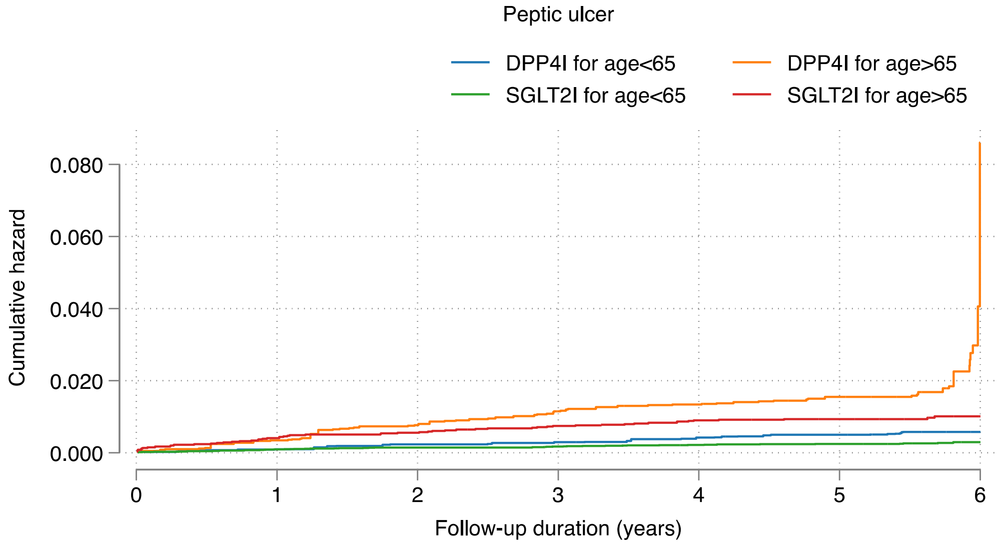

Number at risk

| | 0 | 1 | 2 | 3 | 4 | 5 | 6 |
|---|---|---|---|---|---|---|---|
| DPP4I for age<65 | 17269 | 17252 | 17215 | 17142 | 16972 | 16795 | 90 |
| DPP4I for age>65 | 6173 | 6147 | 6078 | 5933 | 5698 | 5430 | 52 |
| SGLT2I for age<65 | 17489 | 17473 | 17454 | 17414 | 17337 | 17199 | 85 |
| SGLT2I for age>65 | 5953 | 5929 | 5898 | 5834 | 5724 | 5546 | 34 |

Cumulative incidence curve by drug use after propensity score matching

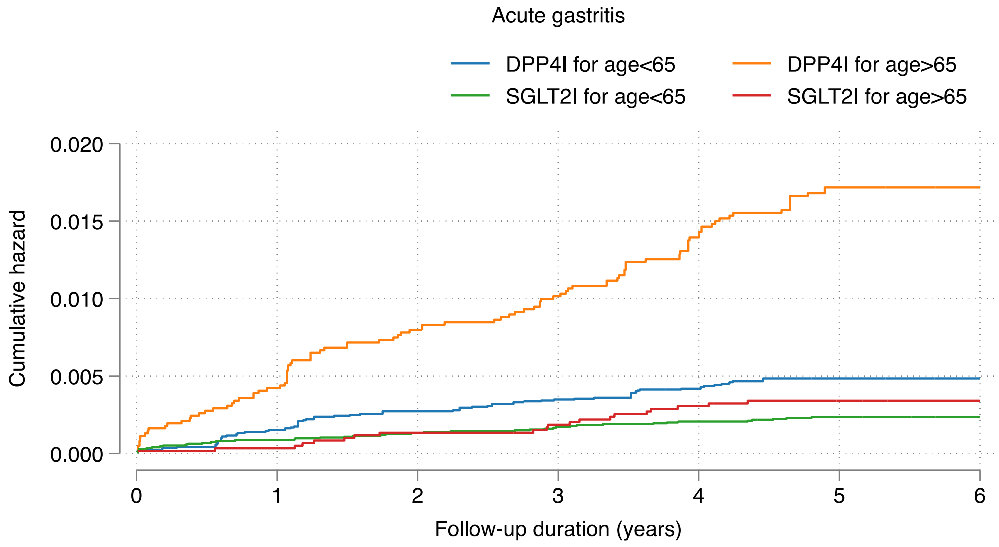

Number at risk

| | 0 | 1 | 2 | 3 | 4 | 5 | 6 |
|---|---|---|---|---|---|---|---|
| DPP4I for age<65 | 17269 | 17242 | 17209 | 17129 | 16966 | 16785 | 90 |
| DPP4I for age>65 | 6173 | 6142 | 6078 | 5932 | 5680 | 5404 | 50 |
| SGLT2I for age<65 | 17489 | 17474 | 17456 | 17412 | 17338 | 17200 | 85 |
| SGLT2I for age>65 | 5953 | 5951 | 5922 | 5866 | 5755 | 5571 | 36 |

Cumulative incidence curve by drug use after propensity score matching

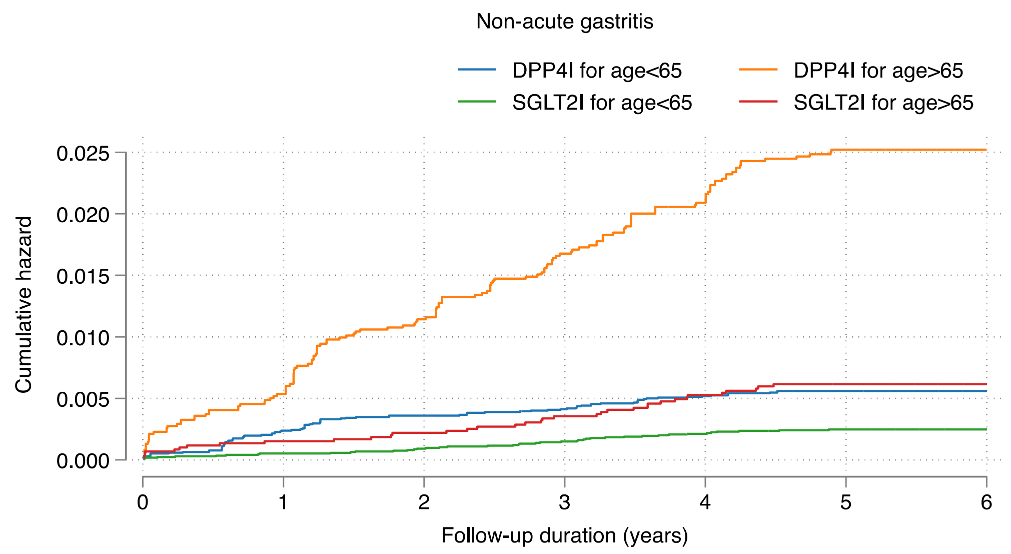

Number at risk

| | | | | | | | |
|---|---|---|---|---|---|---|---|
| DPP4I for age<65 | 17269 | 17227 | 17193 | 17119 | 16948 | 16765 | 90 |
| DPP4I for age>65 | 6173 | 6135 | 6059 | 5895 | 5641 | 5369 | 51 |
| SGLT2I for age<65 | 17489 | 17480 | 17464 | 17418 | 17338 | 17201 | 85 |
| SGLT2I for age>65 | 5953 | 5944 | 5917 | 5858 | 5744 | 5557 | 35 |

Cumulative incidence curve by drug use after propensity score matching

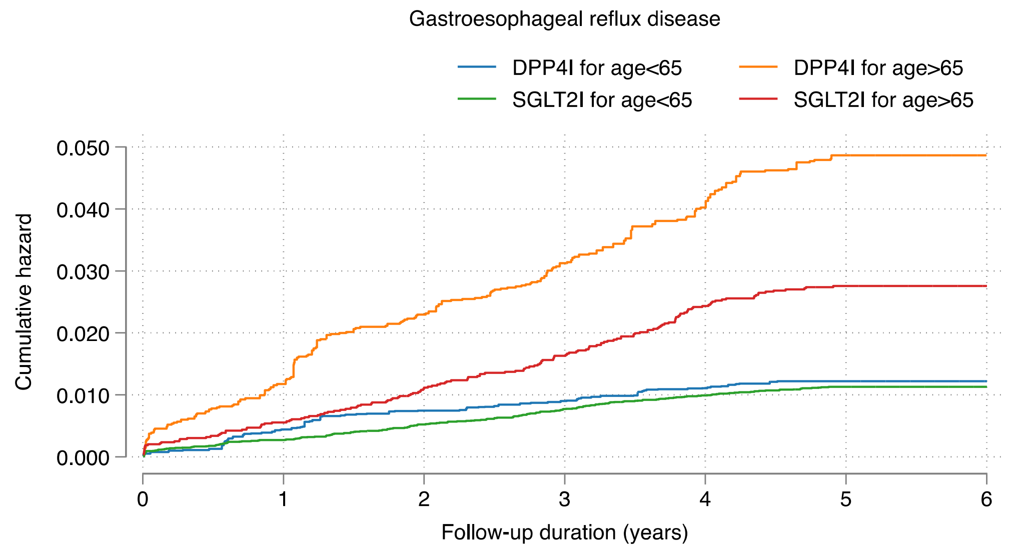

Number at risk

| | | | | | | | |
|---|---|---|---|---|---|---|---|
| DPP4I for age<65 | 17269 | 17192 | 17128 | 17036 | 16858 | 16675 | 90 |
| DPP4I for age>65 | 6173 | 6097 | 5992 | 5817 | 5544 | 5263 | 49 |
| SGLT2I for age<65 | 17489 | 17442 | 17390 | 17312 | 17210 | 17060 | 83 |
| SGLT2I for age>65 | 5953 | 5920 | 5867 | 5787 | 5648 | 5463 | 34 |

Cumulative incidence curve by drug use after propensity score matching

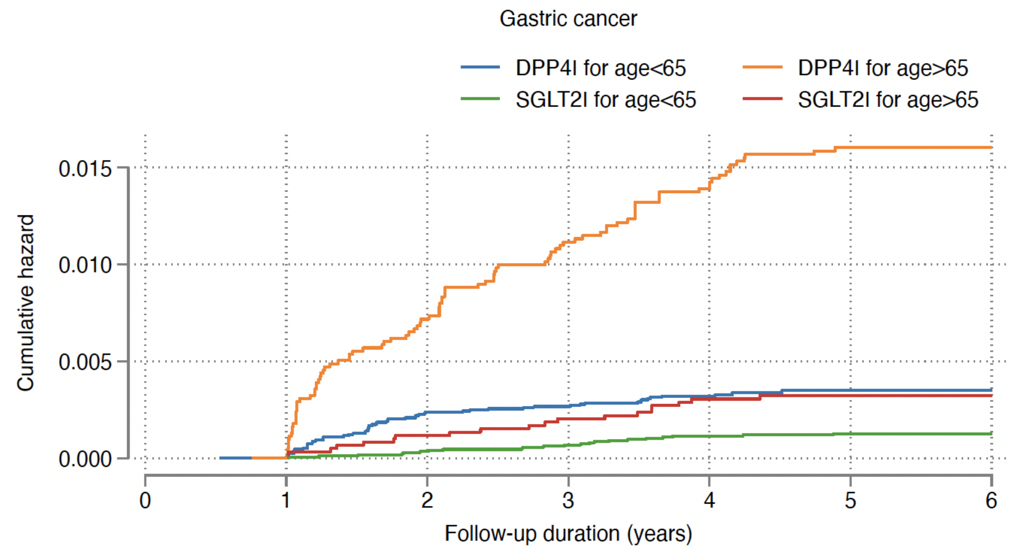

Number at risk

| | | | | | | | |
|---|---|---|---|---|---|---|---|
| DPP4I for age<65 | 17269 | 17268 | 17214 | 17144 | 16980 | 16797 | 90 |
| DPP4I for age>65 | 6173 | 6168 | 6085 | 5926 | 5677 | 5410 | 51 |
| SGLT2I for age<65 | 17489 | 17489 | 17474 | 17431 | 17354 | 17218 | 85 |
| SGLT2I for age>65 | 5953 | 5953 | 5923 | 5865 | 5755 | 5571 | 36 |

Cumulative incidence curve by drug use after propensity score matching

SGLT2I: sodium glucose cotransporter-2 inhibitor; DPP4I: dipeptidyl peptidase-4 inhibitor

**Supplementary Figure 3B. Cumulative incidence curves for new onset gastric outcomes and all-cause mortality stratified by combinations of age and drug exposure effects of SGLT2I and DPP4I in the matched cohort.**

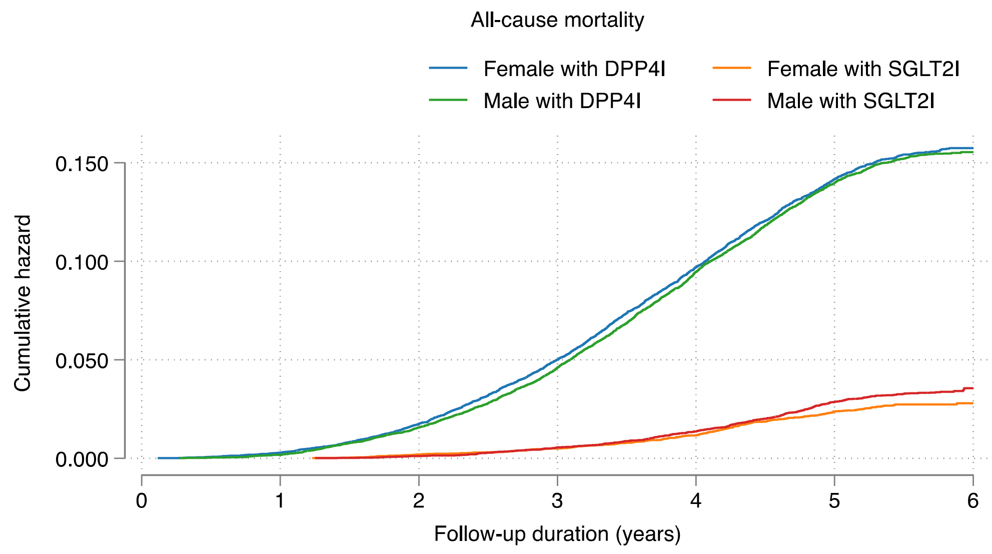

Number at risk

| | | | | | | | |
|---|---|---|---|---|---|---|---|
| Female with DPP4I | 18235 | 18184 | 17919 | 17345 | 16552 | 15826 | 81 |
| Female with SGLT2I | 9463 | 9463 | 9445 | 9416 | 9353 | 9241 | 47 |
| Male with DPP4I | 21181 | 21144 | 20852 | 20231 | 19271 | 18420 | 97 |
| Male with SGLT2I | 13979 | 13979 | 13963 | 13901 | 13790 | 13585 | 74 |

Cumulative incidence curve by drug use before propensity score matching

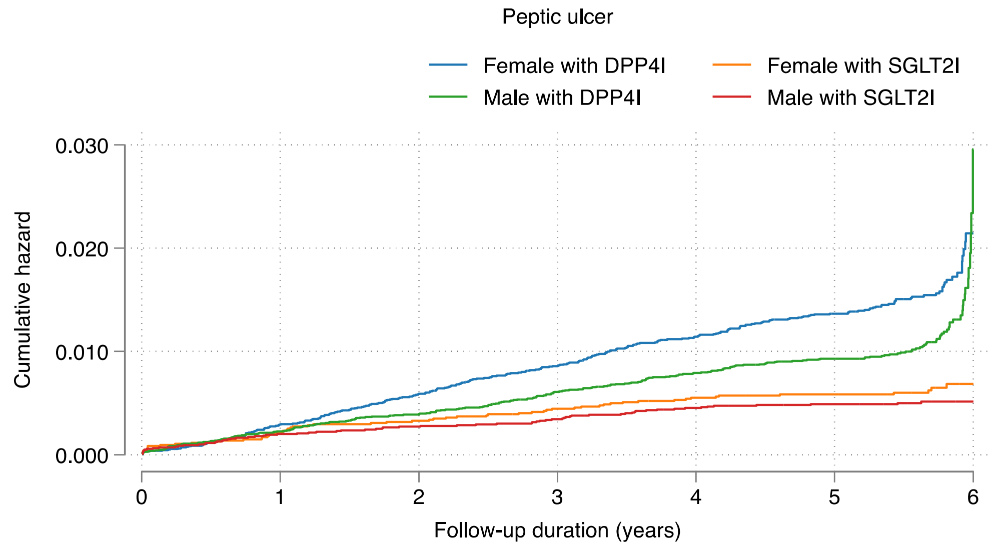

Number at risk

| | | | | | | | |
|---|---|---|---|---|---|---|---|
| Female with DPP4I | 18235 | 18134 | 17829 | 17238 | 16438 | 15716 | 80 |
| Female with SGLT2I | 9463 | 9443 | 9415 | 9375 | 9305 | 9196 | 46 |
| Male with DPP4I | 21181 | 21099 | 20780 | 20135 | 19175 | 18329 | 96 |
| Male with SGLT2I | 13979 | 13951 | 13926 | 13857 | 13733 | 13525 | 71 |

Cumulative incidence curve by drug use before propensity score matching

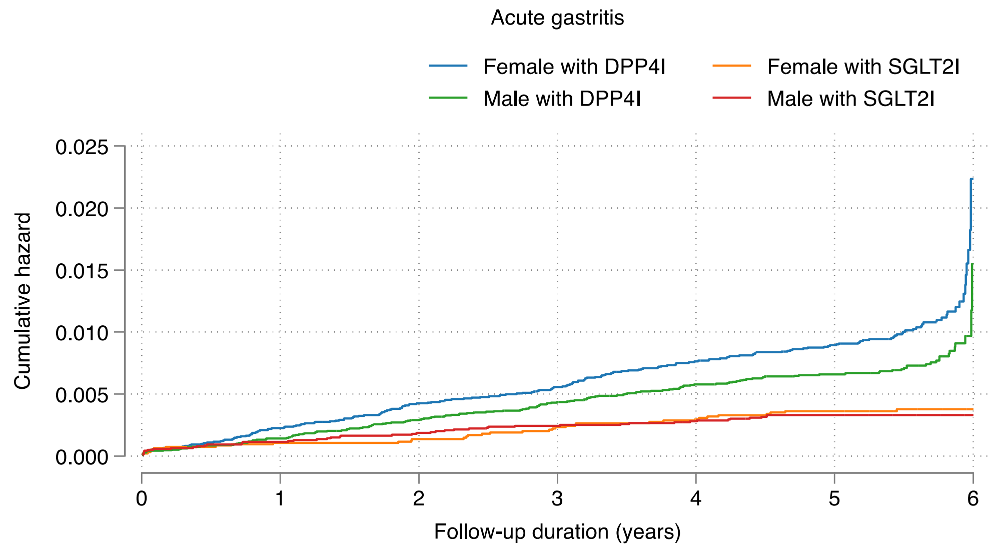

Number at risk

| | | | | | | | |
|---|---|---|---|---|---|---|---|
| Female with DPP4I | 18235 | 18144 | 17853 | 17275 | 16476 | 15751 | 81 |
| Female with SGLT2I | 9463 | 9453 | 9434 | 9396 | 9329 | 9214 | 47 |
| Male with DPP4I | 21181 | 21117 | 20799 | 20166 | 19197 | 18356 | 97 |
| Male with SGLT2I | 13979 | 13963 | 13938 | 13870 | 13756 | 13548 | 73 |

Cumulative incidence curve by drug use before propensity score matching

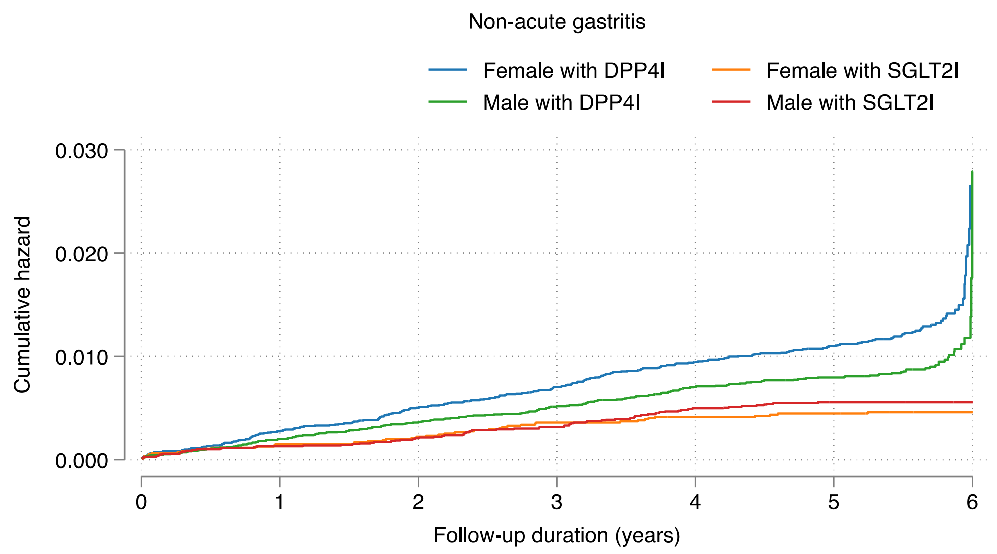

Number at risk

| | | | | | | | |
|---|---|---|---|---|---|---|---|
| Female with DPP4I | 18235 | 18135 | 17839 | 17256 | 16461 | 15739 | 81 |
| Female with SGLT2I | 9463 | 9449 | 9427 | 9385 | 9320 | 9210 | 47 |
| Male with DPP4I | 21181 | 21106 | 20784 | 20151 | 19182 | 18340 | 97 |
| Male with SGLT2I | 13979 | 13961 | 13935 | 13858 | 13724 | 13517 | 71 |

Cumulative incidence curve by drug use before propensity score matching

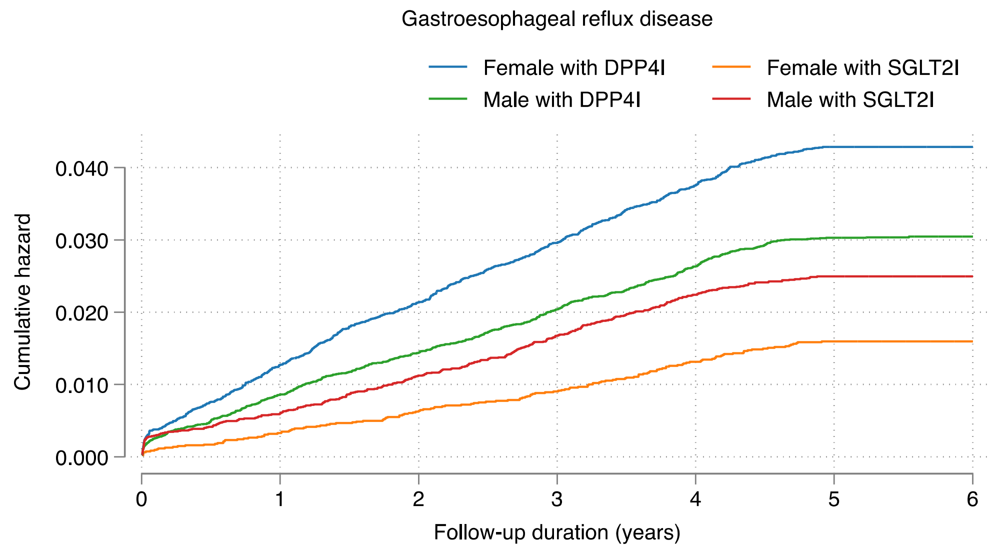

Number at risk

| | | | | | | | |
|---|---|---|---|---|---|---|---|
| Female with DPP4I | 18235 | 17959 | 17577 | 16932 | 16107 | 15388 | 77 |
| Female with SGLT2I | 9463 | 9432 | 9387 | 9333 | 9236 | 9108 | 44 |
| Male with DPP4I | 21181 | 20967 | 20569 | 19883 | 18893 | 18043 | 95 |
| Male with SGLT2I | 13979 | 13896 | 13812 | 13681 | 13511 | 13290 | 73 |

Cumulative incidence curve by drug use before propensity score matching

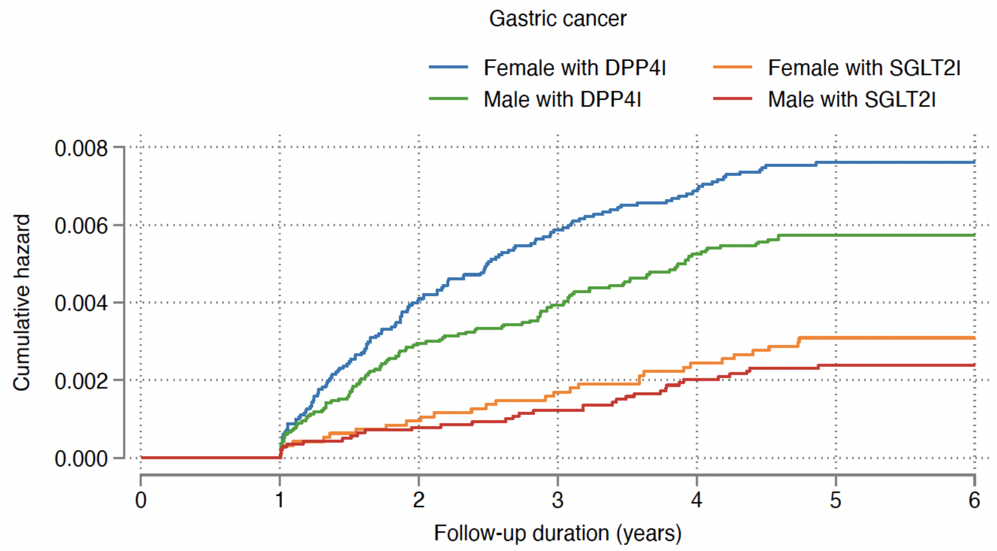

Number at risk

| | | | | | | | |
|---|---|---|---|---|---|---|---|
| Female with DPP4I | 18235 | 18184 | 17849 | 17261 | 16475 | 15748 | 81 |
| Female with SGLT2I | 9463 | 9463 | 9435 | 9395 | 9322 | 9205 | 47 |
| Male with DPP4I | 21181 | 21142 | 20792 | 20164 | 19202 | 18354 | 96 |
| Male with SGLT2I | 13979 | 13967 | 13924 | 13844 | 13717 | 13506 | 74 |

Cumulative incidence curve by drug use before propensity score matching

**Supplementary Figure 4A. Cumulative incidence curves for new onset gastric outcomes and all-cause mortality stratified by combinations of gender and drug exposure effects of SGLT2I and DPP4I before propensity score matching.**

SGLT2I: Sodium-glucose cotransporter-2 inhibitors; DPP4I: Dipeptidyl peptidase-4 inhibitors.

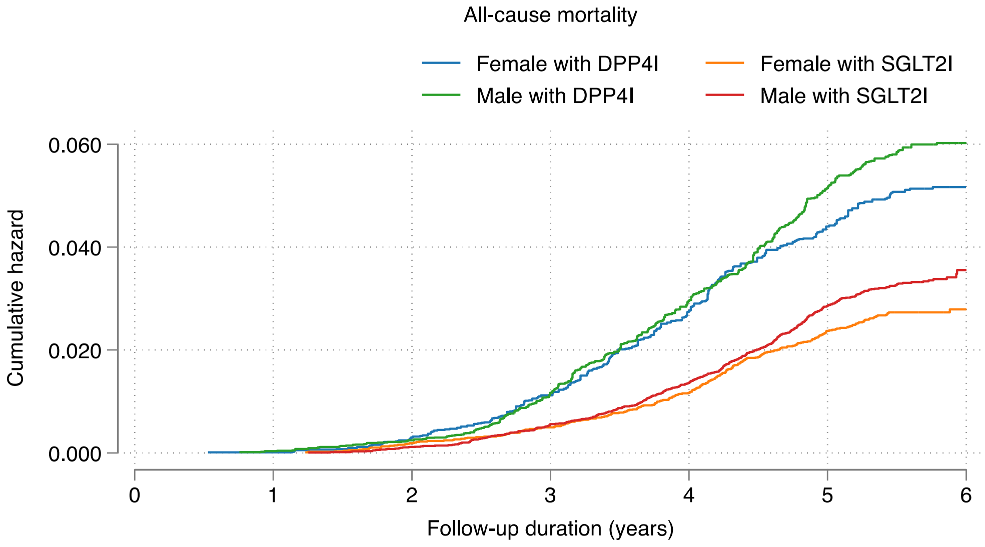

Number at risk

| | | | | | | | |
|---|---|---|---|---|---|---|---|
| Female with DPP4I | 9429 | 9428 | 9401 | 9324 | 9174 | 9023 | 45 |
| Female with SGLT2I | 9463 | 9463 | 9445 | 9416 | 9353 | 9241 | 47 |
| Male with DPP4I | 14013 | 14008 | 13979 | 13850 | 13606 | 13310 | 97 |
| Male with SGLT2I | 13979 | 13979 | 13963 | 13901 | 13790 | 13585 | 74 |

Cumulative incidence curve by drug use after propensity score matching

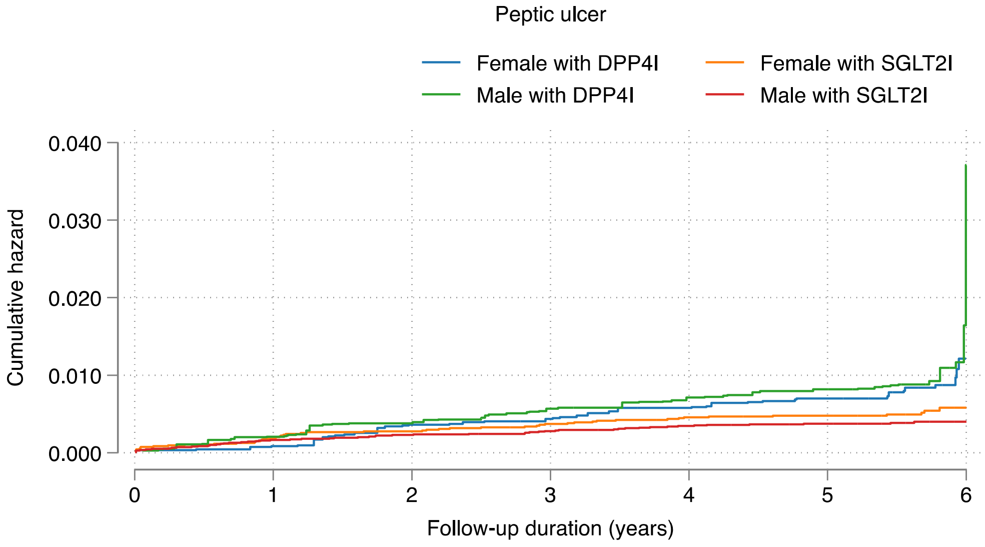

Number at risk

| | | | | | | | |
|---|---|---|---|---|---|---|---|
| Female with DPP4I | 9429 | 9420 | 9367 | 9294 | 9140 | 8988 | 45 |
| Female with SGLT2I | 9463 | 9446 | 9420 | 9382 | 9313 | 9204 | 46 |
| Male with DPP4I | 14013 | 13979 | 13926 | 13781 | 13530 | 13237 | 97 |
| Male with SGLT2I | 13979 | 13956 | 13932 | 13866 | 13748 | 13541 | 73 |

Cumulative incidence curve by drug use after propensity score matching

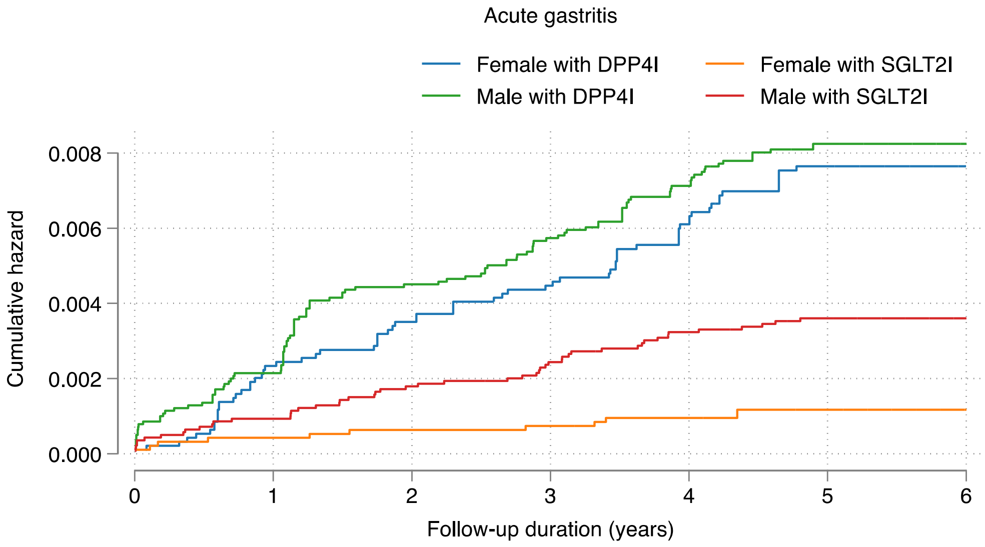

Number at risk

| | | | | | | | |
|---|---|---|---|---|---|---|---|
| Female with DPP4I | 9429 | 9406 | 9370 | 9286 | 9124 | 8963 | 45 |
| Female with SGLT2I | 9463 | 9459 | 9439 | 9409 | 9345 | 9232 | 47 |
| Male with DPP4I | 14013 | 13978 | 13917 | 13775 | 13522 | 13226 | 95 |
| Male with SGLT2I | 13979 | 13966 | 13939 | 13869 | 13748 | 13539 | 74 |

Cumulative incidence curve by drug use after propensity score matching

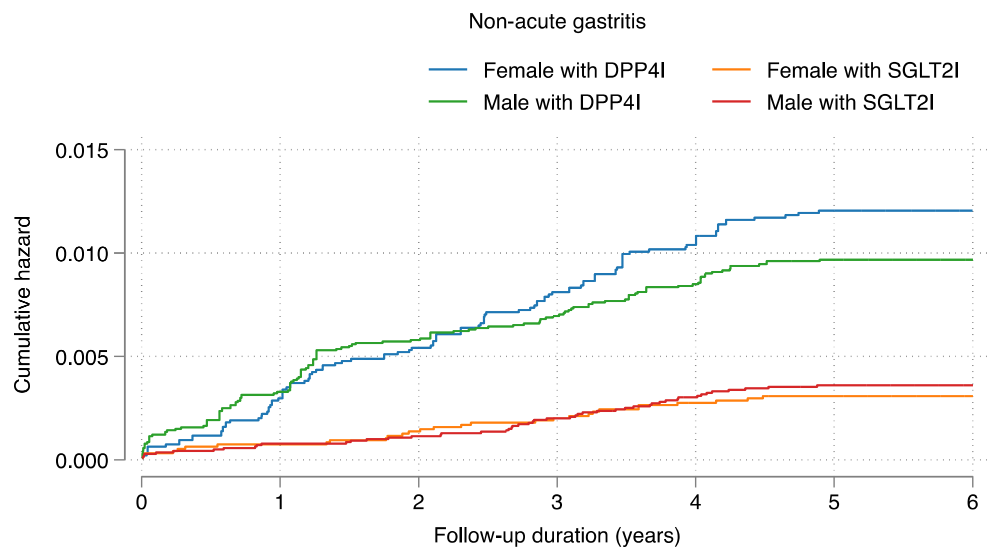

Number at risk

| | | | | | | | |
|---|---|---|---|---|---|---|---|
| Female with DPP4I | 9429 | 9400 | 9354 | 9253 | 9088 | 8938 | 45 |
| Female with SGLT2I | 9463 | 9456 | 9433 | 9399 | 9329 | 9214 | 46 |
| Male with DPP4I | 14013 | 13962 | 13898 | 13761 | 13501 | 13196 | 96 |
| Male with SGLT2I | 13979 | 13968 | 13948 | 13877 | 13753 | 13544 | 74 |

Cumulative incidence curve by drug use after propensity score matching

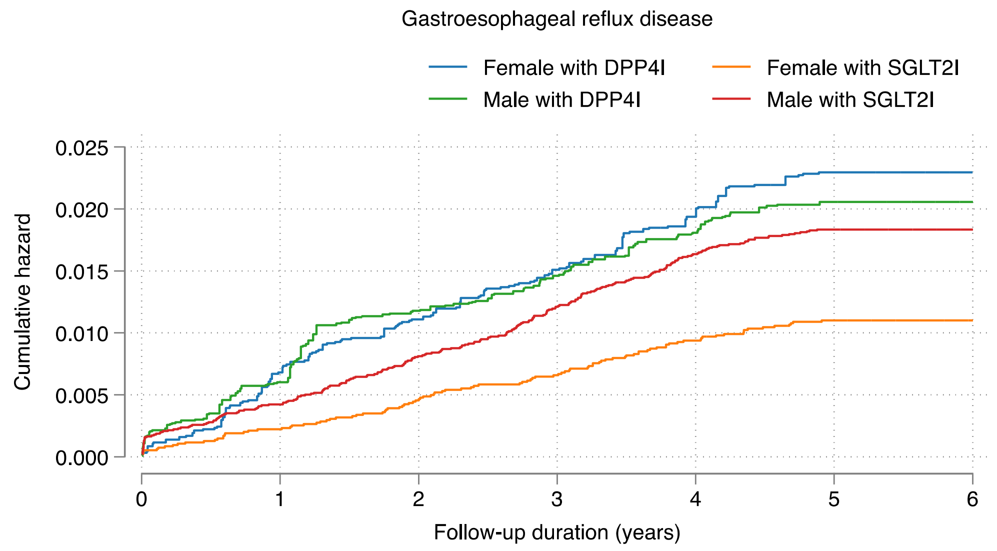

Number at risk

| | | | | | | | |
|---|---|---|---|---|---|---|---|
| Female with DPP4I | 9429 | 9364 | 9303 | 9192 | 9013 | 8853 | 45 |
| Female with SGLT2I | 9463 | 9442 | 9403 | 9356 | 9269 | 9150 | 44 |
| Male with DPP4I | 14013 | 13925 | 13817 | 13661 | 13389 | 13085 | 94 |
| Male with SGLT2I | 13979 | 13920 | 13854 | 13743 | 13589 | 13373 | 73 |

Cumulative incidence curve by drug use after propensity score matching

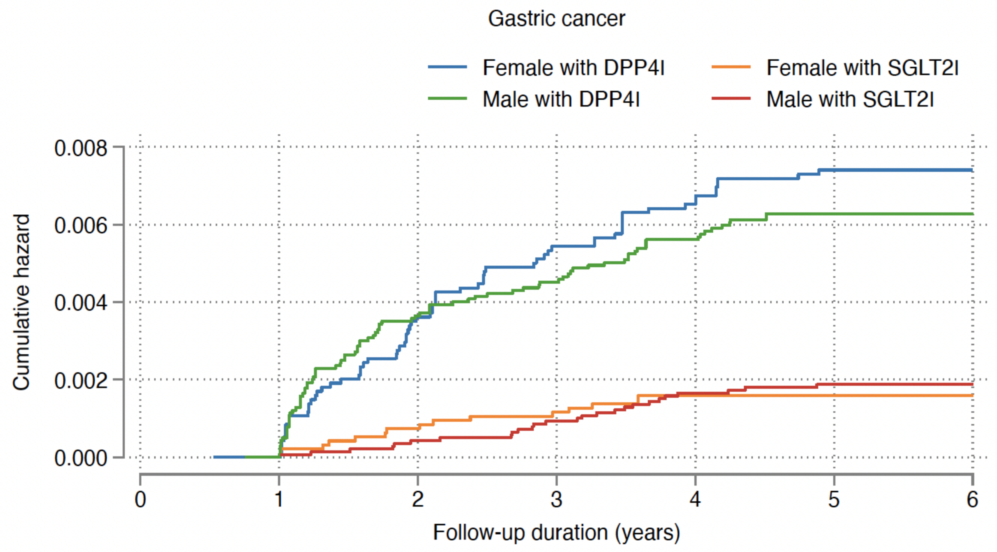

Number at risk

| | | | | | | | |
|---|---|---|---|---|---|---|---|
| Female with DPP4I | 9429 | 9428 | 9371 | 9277 | 9121 | 8972 | 45 |
| Female with SGLT2I | 9463 | 9463 | 9439 | 9406 | 9339 | 9227 | 47 |
| Male with DPP4I | 14013 | 14008 | 13928 | 13793 | 13536 | 13235 | 96 |
| Male with SGLT2I | 13979 | 13979 | 13958 | 13890 | 13770 | 13562 | 74 |

Cumulative incidence curve by drug use after propensity score matching

**Supplementary Figure 4B. Cumulative incidence curves for new onset gastric outcomes and all-cause mortality stratified by combinations of gender and drug exposure effects of SGLT2I and DPP4I in the matched cohort.**

SGLT2I: Sodium-glucose cotransporter-2 inhibitors; DPP4I: Dipeptidyl peptidase-4 inhibitors.

**Supplementary Table 1. The International Classification of Diseases, Clinical Modification (ICD-9-CM) codes for definitions of past comorbidities and outcomes.**

HBV: hepatitis B virus; HCV: hepatitis C virus

| |
|---|
| **Adverse gastric outcomes of interest** |
| **Peptic ulcer:** 531–533 |
| **Acute gastritis:** 535.0 |
| **Non-acute gastritis:** 535.1–535.4, 535.7 |
| **Gastroesophageal reflux disease:** 530.81, 530.11 |
| **Gastric cancer:** 151, 151.0, 151.1, 151.2, 151.3, 151.4, 151.5, 151.6, 151.8, 151.9, 230.2 |
| **Past comorbidities** |
| **Cancer**: 140-239, excluded gastric cancer |
| **Hypertension**: 401 401.1 401.9 402 402.01 402.1 402.11 402.9 402.91 403 403.01 403.1 403.11 403.9 403.91 404 404.01 404.02 404.03 404.1 404.11 404.12 404.13 404.9 404.91 404.92 404.93 405 405.01 405.09 405.1 405.11 405.19 405.9 405.91 405.99 437.2 |
| **Chronic liver disease and cirrhosis**: 571.0 571.1 571.2 571.3 571.40 571.41 571.42 471.49 571.5 571.6 571.8, 571.9 |
| **HBV infection:** 070.20–070.23, 070.30–070.33, V02.61 |
| **HCV infection:** 070.41, 070.44, 070.51, 070.54, 070.7, 070.70, 070.71 |
| **History of acute liver injury:** 570 572.2 |
| **Other liver disease:** 275.1 275.0 572.0 572.4 572.1 572.3 572.8 573.0 573.4 573.8 573.9 |
| **Autoimmune diseases:** 136.1, 359.79, 359.71, 443.1, 446, 555, 556.8~556.9, 556.0~556.6, 695.4, 710, 714, 720, 725 |
| **Polyps:** 569.0, 211.3 |
| **Gastrointestinal bleeding:** 578.9 |
| **Heart failure:** 428 428 428.1 428.2 428.2 428.21 428.22 428.23 428.3 428.3 428.31 428.32 428.33 428.4 428.4 428.41 428.42 428.43 428.9 398.91 402.01 402.11 402.91 404.01 404.03 404.11 404.13 404.91 404.93 |
| **Atrial fibrillation:** 427.31 429.4 |
| **Stroke/transient ischemic attack:** 435 435.1 435.2 435.3 435.8 435.9 433.81 433.91 434 436 437 437.1 433.31 433.01 434.01 434.1 434.11 434.9 434.91 437.2 437.3 437.4 437.5 437.6 437.7 437.8 437.9 430 431 432 432.1 432.9 |
| **Ischemic heart disease:** 410.01 410.02 410.1 410.11 410.12 410.2 410.21 410.22 410.3 410.31 410.32 410.4 410.41 410.42 410.5 410.51 410.52 410.6 410.61 410.62 410.7 410.71 410.72 410.8 410.81 410.82 410.9 410.91 410.92 411 411.1 411.8 411.81 411.89 413 413.1 413.9 414 414.01 414.02 414.03 414.04 414.05 414.06 414.07 414.1 414.11 414.12 414.19 414.2 414.3 414.4 414.8 414.9 410 412 |
| **Peripheral vascular disease:** 250.7 443.9 443 443.1 443.2 443.21 443.22 443.23 443.24 443.29 443.8 443.81 443.82 443.89 441 443.9 785.4 V43.4 |

| |
|---|
| **Acute myocardial infarction:** 410 410.01 410.02 410.1 410.11 410.12 410.2 410.21 410.22 410.3 410.31 410.32 410.4 410.41 410.42 410.5 410.51 410.52 410.6 410.61 410.62 410.7 410.71 410.72 410.8 410.81 410.82 410.9 410.91 410.92 |
| **H. pylori infection:** 041.86 (ICD-9 codes) + MIS data + H. pylori eradication therapy |
| **Renal diseases:** 582 582.1 582.2 582.4 582.8 582.81 582.89 582.9 583 583 583.1 583.2 583.4 583.6 583.7 585 585.1 585.2 585.3 585.4 585.5 585.6 585.9 586 588 588 588.1 588.8 588.81 588.89 588.9 |

**Supplementary Table 2. Calculations for variability measure**

| Variability measure | Definition |
|---|---|
| Standard deviation (SD) | $\sqrt{\frac{\sum_{i=1}^{Number\ of\ measurements}(test_i - individual\ mean)^2}{Number\ of\ measurements}}$ |
| Variance | $\frac{\sum_{i=1}^{Number\ of\ measurements}(test_i - individual\ mean)^2}{Number\ of\ measurements}$ |
| Standard deviation/Initial (SD/initial) | $\frac{SD}{individual\ initial\ value}$ |
| Coefficient of variation (CV) | $\frac{SD}{individual\ mean}$ |

.

**Supplementary Table 3. Multivariate Cox regression models with adjustments to predict new gastric cancer and gastric diseases in the SGLT2I v.s. DPP4I matched cohort.**

* for p≤ 0.05, ** for p ≤ 0.01, *** for p ≤ 0.001; HR: hazard ratio; CI: confidence interval; SGLT2I: sodium glucose cotransporter-2 inhibitor; DPP4I: dipeptidyl peptidase-4 inhibitor.

Model 1 adjusted for significant demographics.

Model 2 adjusted for significant demographics, and past comorbidities.

Model 3 adjusted for significant demographics, past comorbidities, and non-SGLT2I/DPP4I medications.

Model 4 adjusted for significant demographics, past comorbidities, non-SGLT2I/DPP4I medications, abbreviated MDRD, and NLR

Model 5 adjusted for significant demographics, past comorbidities, non-SGLT2I/DPP4I medications, abbreviated MDRD, NLR, fasting glucose, HbA1c, and duration from earliest diabetes mellitus date to initial drug exposure date.

| **Characteristics** | **Peptic ulcer HR [95% CI];P value** | **Acute gastritis HR [95% CI];P value** | **Non acute gastritis HR [95% CI];P value** | **Gastroesophageal reflux disease HR [95% CI];P value** | **Gastric cancer HR [95% CI];P value** | **All-cause mortality HR [95% CI];P value** |
|---|---|---|---|---|---|---|
| ***Model 1*** | 0.50[0.39-0.63];<0.0001*** | 0.33[0.25-0.45];<0.0001*** | 0.33[0.25-0.42];<0.0001*** | 0.73[0.64-0.83];<0.0001*** | 0.27[0.19-0.38];<0.0001*** | 0.56[0.51-0.62];<0.0001*** |
| ***Model 2*** | 0.49[0.38-0.62];<0.0001*** | 0.33[0.25-0.44];<0.0001*** | 0.32[0.25-0.41];<0.0001*** | 0.71[0.62-0.81];<0.0001*** | 0.26[0.18-0.37];<0.0001*** | 0.55[0.50-0.60];<0.0001*** |
| ***Model 3*** | 0.534[0.419-0.680];<0.0001*** | 0.387[0.288-0.520];<0.0001*** | 0.342[0.264-0.443];<0.0001*** | 0.798[0.693-0.918];0.0016** | 0.277[0.195-0.392];<0.0001*** | 0.63[0.57-0.69];<0.0001*** |
| ***Model 4*** | 0.58[0.43-0.78];0.0003*** | 0.30[0.20-0.44];<0.0001*** | 0.32[0.23-0.43];<0.0001*** | 0.60[0.50-0.72];<0.0001*** | 0.27[0.18-0.41];<0.0001*** | 0.73[0.64-0.83];<0.0001*** |
| ***Model 5*** | 0.66[0.47-0.91];0.0118* | 0.31[0.20-0.48];<0.0001*** | 0.35[0.25-0.49];<0.0001*** | 0.62[0.50-0.76];<0.0001*** | 0.30[0.19-0.48];<0.0001*** | 0.77[0.66-0.89];0.0003*** |

**Supplementary Table 4. Sensitivity analysis: Three-arm (only SGLT2I, only DPP4I, and only GLP1a) analysis results using stabilized IPTW**

The following exclusion criteria applied

1. With prior peptic ulcer, gastritis, gastroesophageal reflux disease, and gastric cancer (N=108)
2. Patients who died within 30 days after initial drug exposure (N=167)
3. Without complete demographics (N=19)
4. Under 18 years old (N=108)
5. Used at least two drugs from SGLT21, DPP4I, and GLP1a (N=12956)
6. New onset gastric cancer development less than 1 year after drug exposure (N=29)

* for p≤ 0.05, ** for p ≤ 0.01, *** for p ≤ 0.001; HR: hazard ratio; CI: confidence interval; SGLT2I: sodium glucose cotransporter-2 inhibitor; DPP4I: dipeptidyl peptidase-4 inhibitor.

| Model | Peptic ulcer HR [95% CI];P value | Acute gastritis HR [95% CI];P value | Non acute gastritis HR [95% CI];P value | Gastroesophageal reflux disease HR [95% CI];P value | Gastric cancer HR [95% CI];P value | All-cause mortality HR [95% CI];P value |
|---|---|---|---|---|---|---|
| DPP4I v.s. SGLT2I | 2.19[1.76-2.72];<0.0001*** | 3.06[2.33-4.01];<0.0001*** | 2.25[1.89-2.68];<0.0001*** | 1.58[1.40-1.78];<0.0001*** | 2.18[1.69-2.80];<0.0001*** | 2.57[2.36-2.79];<0.0001*** |
| GLP1a v.s. SGLT2I | 1.57[0.80-3.05];=0.1864 | 1.03[0.49-2.19];=0.9343 | 1.51[0.99-2.32];=0.5818 | 1.43[1.04-1.97];=0.0299* | 2.47[1.27-4.81];=0.0079** | 0.84[0.61-1.15];= 0.2659 |

**Supplementary Table 5. Sensitivity analyses: Exposure effects of SGLT2I v.s. DPP4I on new onset gastric diseases in the SGLT2I v.s. DPP4I matched cohort across different hazard models and matching methods.**

* for p≤ 0.05, ** for p ≤ 0.01, *** for p ≤ 0.001; SGLT2I: Sodium-glucose cotransporter-2 inhibitors; DPP4I: Dipeptidyl peptidase-4 inhibitors; HR: hazard ratio; CI: confidence interval

| Model | Peptic ulcer HR [95% CI];P value | Acute gastritis HR [95% CI];P value | Non acute gastritis HR [95% CI];P value | Gastroesophageal reflux disease HR [95% CI];P value | Gastric cancer HR [95% CI];P value | All-cause mortality HR [95% CI];P value |
|---|---|---|---|---|---|---|
| Cause-specific hazard models | 0.52[0.41-0.67];<0.0001*** | 0.354[0.265-0.474];<0.0001*** | 0.348[0.270-0.449];<0.0001*** | 0.756[0.659-0.867];0.0001*** | 0.282[0.199-0.398];<0.0001*** | 0.60[0.55-0.66];<0.0001*** |
| Sub-distribution hazard models | 0.49[0.38-0.62];<0.0001*** | 0.33[0.24-0.44];<0.0001*** | 0.32[0.25-0.41];<0.0001*** | 0.71[0.62-0.82];<0.0001*** | 0.26[0.19-0.37];<0.0001*** | 0.55[0.50-0.60];<0.0001*** |
| PS stratification | 0.66[0.47-0.91];0.0124* | 0.31[0.20-0.49];<0.0001*** | 0.34[0.24-0.48];<0.0001*** | 0.62[0.50-0.76];<0.0001*** | 0.30[0.19-0.47];<0.0001*** | 0.77[0.66-0.89];0.0003*** |
| PS with IPTW | 0.52[0.40-0.67];<0.0001*** | 0.35[0.25-0.48];<0.0001*** | 0.33[0.26-0.44];<0.0001*** | 0.67[0.57-0.78];<0.0001*** | 0.27[0.19-0.39];<0.0001*** | 0.64[0.58-0.72];<0.0001*** |
| PS with SIPTW | 0.48[0.38-0.61];<0.0001*** | 0.33[0.24-0.43];<0.0001*** | 0.32[0.25-0.41];<0.0001*** | 0.71[0.62-0.81];<0.0001*** | 0.26[0.18-0.37];<0.0001*** | 0.55[0.50-0.60];<0.0001*** |

**Supplementary Table 6. Sensitivity analysis: Excluding patients with CKD stage 4/5 (eGFR <30), peritoneal dialysis or haemodialysis in the SGLT2I v.s. DPP4I matched cohort.**

* for p≤ 0.05, ** for p ≤ 0.01, *** for p ≤ 0.001; SGLT2I: Sodium-glucose cotransporter-2 inhibitors; DPP4I: Dipeptidyl peptidase-4 inhibitors; HR: hazard ratio; CI: confidence interval

| | Peptic ulcer HR [95% CI];P value | Acute gastritis HR [95% CI];P value | Non-acute gastritis HR [95% CI];P value | Gastroesophageal reflux disease HR [95% CI];P value | Gastric cancer HR [95% CI];P value | All-cause mortality HR [95% CI];P value |
|---|---|---|---|---|---|---|
| SGLT2I v.s. DPP4I | 0.45[0.35-0.58];<0.0001*** | 0.29[0.21-0.40];<0.0001*** | 0.31[0.24-0.40];<0.0001*** | 0.59[0.51-0.69];<0.0001*** | 0.25[0.18-0.36];<0.0001*** | 0.55[0.50-0.61];<0.0001*** |

**Supplementary Table 7. Sensitivity analysis: Consideration of 1-year lag time effects in the SGLT2I v.s. DPP4I matched cohort.**

* for p≤ 0.05, ** for p ≤ 0.01, *** for p ≤ 0.001; SGLT2I: Sodium-glucose cotransporter-2 inhibitors; DPP4I: Dipeptidyl peptidase-4 inhibitors; HR: hazard ratio; CI: confidence interval

| | Peptic ulcer HR [95% CI];P value | Acute gastritis HR [95% CI];P value | Non-acute gastritis HR [95% CI];P value | Gastroesophageal reflux disease HR [95% CI];P value | Gastric cancer HR [95% CI];P value | All-cause mortality HR [95% CI];P value |
|---|---|---|---|---|---|---|
| SGLT2I v.s. DPP4I | 0.36[0.27-0.48];<0.0001*** | 0.33[0.23-0.46];<0.0001*** | 0.35[0.26-0.47];<0.0001*** | 0.79[0.67-0.92];0.0028** | 0.26[0.18-0.37];<0.0001*** | 0.55[0.50-0.61];<0.0001*** |

**Supplementary Table 8. Sensitivity analysis: As-treated approach of SGLT2I v.s. DPP4I on new onset gastric diseases in the SGLT2I v.s. DPP4I matched cohort**

| | Peptic ulcer HR [95% CI];P value | Acute gastritis HR [95% CI];P value | Non-acute gastritis HR [95% CI];P value | Gastroesophageal reflux disease HR [95% CI];P value | Gastric cancer HR [95% CI];P value | All-cause mortality HR [95% CI];P value |
|---|---|---|---|---|---|---|
| SGLT2I v.s. DPP4I | 0.40[0.33-0.48];<0.0001*** | 0.23[0.18-0.29];<0.0001*** | 0.32[0.28-0.38];<0.0001*** | 0.57[0.52-0.64];<0.0001*** | 0.28[0.23-0.36];<0.0001*** | 0.19[0.18-0.21];<0.0001*** |
